# Neural characterisation of persistent quantitative and qualitative COVID-19-related olfactory dysfunction: The COVORTS study

**DOI:** 10.64898/2026.09.07.26362431

**Authors:** Birgit van Dijk, Elbrich M. Postma, Louise M. Leenders, Puck Smits, Digna M.A. Kamalski, Paul A.M. Smeets, Sanne Boesveldt

## Abstract

The mechanism underlying persistent COVID-19-related olfactory dysfunction (OD) remains unknown. There are indications for both peripheral (within the nose) and central (in the brain) mechanisms. Patients can experience a decrease in olfactory ability (quantitative OD) or a distorted odour perception (qualitative OD). The mechanisms might differ between these two symptom presentations. In this (f)MRI study, we investigated structural and functional brain alterations in patients with persistent (> 1 month) quantitative OD (Sniffin’ Sticks score < 30.75, n=21) or parosmia (self-reported, n=27) following SARS-CoV-2 infection, aged 19-60 years and, on average, included 472 days after diagnosis (the COVORTS study). Additionally, a control group of 49 normosmic subjects aged 22-60 years was included. Olfactory bulb volume (OBV) was determined by manual segmentation; whole-brain and regional grey matter density (GMD) of olfactory-related areas was assessed using voxel-based morphometry; and odour-induced brain activation was examined with an olfactory fMRI task. When grouped together, patients with OD had a smaller OBV than controls (84.4 vs 95.3 mm3, *p*=0.026). The two patient groups did not differ in OBV. ROI analysis of olfactory-related brain regions at an exploratory threshold showed lower GMD in primary and secondary olfactory regions in patients compared to controls, which appears to be mainly driven by quantitative OD. No difference in odour-induced brain activation was found. The observed smaller olfactory bulbs, along with modest findings beyond this primary olfactory structure, provide important insights into potential mechanisms of persistent COVID-19-related OD and highlight the need for longer follow-up to evaluate further neural degradation or recovery.

## 1. Introduction

Over six years after the onset of the SARS-CoV-2 pandemic, many people still experience its long-term effects. Post-COVID syndrome is defined by patients experiencing symptoms for over three months after initial infection [1]. Reported prevalence varies widely, from 15-78% depending on the definition used and elapsed time since infection onset [2–6], and appears to be less common following the Omicron strain of the virus [7]. Commonly reported symptoms of post-COVID syndrome are shortness of breath, fatigue, cognitive and memory issues, joint and muscle pain and high heart rate [3, 8]. Another frequently reported long-term consequence of COVID-19 infection is olfactory dysfunction (OD) [9]. While a disordered sense of smell may seem like a minor symptom in the context of post-COVID syndrome, the ramifications for patient well-being can be far-reaching. Persistent acquired OD significantly affects patients’ quality of life and eating behaviour [10, 11] and is associated with depression [12], suicidal ideation [13, 14], and various forms of cognitive deterioration [15].

Transient symptoms – experienced by the vast majority of patients with COVID-19-related olfactory dysfunction (C19OD) – often involve the sudden onset of a complete loss of smell function followed by a quick recovery (within one to three weeks) [16]. The cause of this transient loss is now generally accepted to involve infection of and damage to the sustentacular support cells within the olfactory epithelium [16, 17]. This damage disrupts the functioning of olfactory receptor neurons, which recover quickly due to the quick turnover rate of sustentacular cells. However, the mechanism behind persistent OD – experienced by around 5% of infected patients [18] – is still poorly understood, resulting in a lack of adequate treatment for the millions of patients experiencing these symptoms worldwide [19, 20].

Whereas patients experiencing transient olfactory loss generally have a similar symptom pattern involving the sudden and complete loss of olfaction with a quick recovery [16], patients with persistent C19OD present with a more variable symptom progression. For these patients, symptoms also tend to start with an acute and complete loss of quantitative olfactory function (i.e., anosmia) [9, 21], which, for most, develops into a decreased sense of smell (i.e., hyposmia) over time [22–24]. A substantial number of patients also experience qualitative OD, especially a distorted sense of smell (i.e., parosmia), which can appear as a solitary symptom or in concurrence with quantitative dysfunction [24–26]. Furthermore, fluctuations in the prevalence of both quantitative and qualitative symptoms have been described [26, 27].

Both acute and persistent quantitative and qualitative OD were well-known – although poorly understood – consequences of pre-pandemic viral upper respiratory tract infections. Therefore, the proposed hypotheses behind the mechanism of persistent symptoms for both pre-pandemic post-viral and C19OD are similar and involve issues within the periphery (the nose) or central alterations (the brain) [16, 17]. Regarding quantitative dysfunction, hypotheses involve the chronic inflammation of the olfactory epithelium resulting in dysfunction of olfactory receptor neurons, followed by a loss of signal transfer between the nose and olfactory bulbs [16, 17]. For parosmia, the main form of qualitative OD, proposed mechanisms involve possible aberrant rewiring of glomeruli into the olfactory bulb after regeneration of the olfactory receptor neurons, or the abnormal or incomplete activation of receptors due to partial damage [16, 28]. Furthermore, both quantitative and qualitative OD could be due to structural or functional alterations in the primary olfactory cortex and related cortical areas [16, 17, 28].

Several magnetic resonance imaging (MRI) studies exploring potential mechanisms underlying persistent C19OD have reported smaller olfactory bulb volumes [29–31], which may indicate decreased signal transfer from the periphery [32] or direct damage to the bulbs themselves [17]. However, (f)MRI studies have also found structural and functional alterations in the olfactory cortices [30, 33–37]. Most studies investigating short- or long-term C19OD focus solely on (severe) quantitative symptoms or do not differentiate between patients with quantitative and qualitative symptoms. This limits the exploration of a possible divergent mechanism of the two symptom presentations. Furthermore, investigations into both structural and functional alterations in the same group of patients are sparse, resulting in a more limited perspective on the pathophysiology of persistent symptoms.

The current study aimed to further elucidate the mechanism underlying persistent COVID-19-related olfactory dysfunction. The *COVid cohORT on Smell loss* (COVORTS) study consists of a longitudinal follow-up of patients with persistent C19OD. The study investigated the course of persistent COVID-19-related chemosensory dysfunction (see [26, 38]) and its consequences on patient well-being (see [39]). Moreover, the cohort includes a sub-population in which (f)MRI measurements have been performed. Using structural and functional MRI, we examined olfactory bulb volume (OBV), grey matter density (GMD), and odour-induced brain activation (BOLD) in patients from the COVORTS study with persistent COVID-19-related quantitative OD or parosmia, and in a control group of healthy (normosmic) individuals.

## 2. Methods

This manuscript uses data from two cohort studies, both of which were conducted according to the Declaration of Helsinki and approved by an accredited medical research ethics committee (MREC Oost-Nederland; https://onderzoekmetmensen.nl/nl/trial/54465 & https://onderzoekmetmensen.nl/nl/trial/49925).

### 2.1. Participants

#### 2.1.1. COVID-19 patients

The patient group in this study was part of the COVORTS (COVid cohORT on Smell loss) study. This cohort study includes a longitudinal follow-up of OD symptoms and their consequences in patients with persistent C19OD, with a sub-population including MRI measurements. Results from this cohort study, including longitudinal prevalence and recovery rate of olfactory, gustatory and trigeminal function and quality of life and appetite and hunger ratings, have previously been published [26, 38, 39]. The COVORTS study included patients aged 18 to 60 years with persistent (> 1 month) self-reported OD due to COVID-19 infection, confirmed by a positive PCR test or a positive SARS-CoV-2 Antigen self-test. Patients were excluded if they had a pre-existing smell and/or taste disorder, were pregnant/intended to become pregnant within the duration of study participation, or were normosmic according to the psychophysical olfaction test performed during screening without having co-occurring self-reported parosmia complaints (Sniffin’ Sticks test score < 30.75 [40]; “normosmic” test scores of patients with self-reported parosmia complaints were not considered for exclusion; measurement of parosmia symptoms is explained in section 2.2.3.). For this MRI sub-study, patients were also excluded if they suffered from claustrophobia or had other MRI contraindications, or had a visual impairment that could not be corrected with MRI-safe glasses or contact lenses. The majority of patients (n=29) in the current manuscript were included through their participation in the longitudinal COVORTS study. A smaller group of patients (n=19) only participated in the MRI sub-study. Recruitment tactics for the COVORTS study have been previously reported [26, 38, 39]. The patients who participated only in the MRI sub-study were recruited using the same strategies, including recontacting people who had applied for the longitudinal COVORTS study but did not meet the stricter inclusion criteria surrounding the time since SARS-CoV-2 diagnosis.

An overview of the inclusion process of the patient group is shown in Fig. 1. The final 48 patients in the current analysis were diagnosed with SARS-CoV-2 between April 2020 and February 2023 and scanned between January 2022 and June 2024 (more information on days since SARS-CoV-2 diagnosis and inclusion can be found in Table 1 in the results section).

**Fig. 1.**
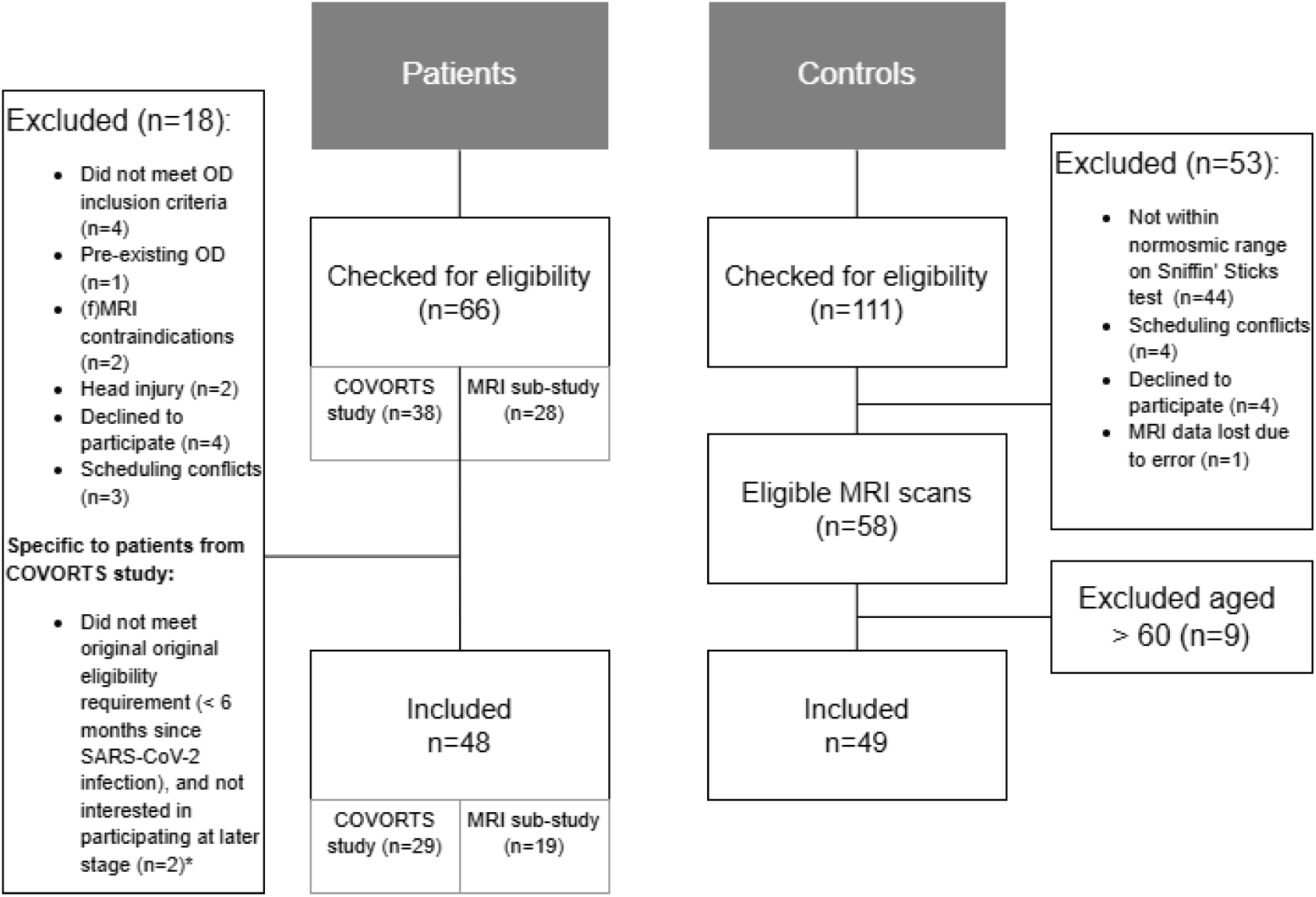
Inclusion flowchart of patients and controls. *In an earlier version of the study protocol, patients had to be within six months of their SARS-CoV-2 infection to participate. Due to scheduling conflicts, these patients from the longitudinal COVORTS study were unable to undergo an MRI session before this six-month criterion had elapsed. These patients were contacted again after this inclusion criterion was dropped.

#### 2.1.2. Normosmic control participants

The control participants were part of the Smell it! study that examined brain morphology and activation in response to odour stimulation in people with normal olfactory function. This study aimed to establish a sex- and age-balanced control group of participants aged between 18-75 years with a self-reported normal sense of smell (normosmic), which was confirmed through psychophysical testing of olfactory functioning (Sniffin’ Sticks test score ≥ 30.75 [40]). Exclusion criteria based on claustrophobia, MRI contraindications and eyesight were the same as for the patient group. Participants were recruited through traditional and social media, the researchers’ own networks and an existing internal participant database. The full Smell it! study cohort consists of 58 participants. For the current analysis, controls older than 60 years were excluded to better match the age distribution of the patient sample, resulting in 49 normosmic controls aged between 19 and 60 years. These patients were scanned between February 2022 and May 2024. An overview of the inclusion process of the control group is shown in Fig. 1.

All participants provided written informed consent and received financial compensation for taking part in the study.

### 2.2. Study procedures

#### 2.2.1. Screening

Interested participants first filled out an online questionnaire to check their eligibility. Upon meeting the study criteria, patients and controls were screened on olfactory functioning, and patients were asked about the presence of parosmia symptoms (see 2.2.2 and 2.2.3 for details). The screening session for both patients and controls also included the Taste Strips test [41], and patients underwent further extensive assessment of chemosensory functioning (as described in Boesveldt et al. [38], van Dijk et al. [26], and van Dijk et al. [39]). The results of the Taste Strips test and further patient assessment are not included in the present manuscript. The majority of patients (n=29) were tested at home during home visits conducted as part of the ongoing longitudinal COVORTS study. For this group of patients, the measurements from the home visit preceding their respective MRI session were used.

The smaller group of patients (n=19) who only participated in the MRI sub-study followed a shortened screening procedure (excluding the home-use test; see [39]), which was performed either at home (n=1) or on the research premises (n=18). The majority of controls were tested on the research premises (n=46), with a few undergoing screening at home for convenience (n=3). After meeting the eligibility criteria, patients and controls who were screened on the research premises underwent a familiarisation session in a mock MRI scanner. This session lasted around 5 minutes, during which participants were exposed to the scan setup and sounds, and performed a short version of the experimental fMRI task. The three controls who were screened at their homes had previous experience with a (clinical) MRI scan. As the fMRI task began with a set of warm-up questions to familiarise participants with using the button box to answer questions (see section 2.2.5 for more details), the familiarisation session was deemed unnecessary for this group of control participants. The patients from the longitudinal COVORTS study were familiar with the research team, so we decided that a familiarisation session was unnecessary.

#### 2.2.2. Assessment of psychophysical olfactory function

Olfactory functioning of both patients and controls was measured using the Sniffin’ Sticks test battery (Burghart Messtechnik, Holm, Germany) [42]. This test battery assesses odour threshold, odour discrimination, and odour identification. For the patient groups, the extended version of the Sniffin’ Sticks identification test was used as an additional measure to mitigate learning effects given the longitudinal nature of the COVORTS study. This extended version contains 32 odours instead of 16 for the identification test [43]. During the identification task, still only 16 of these are presented, randomised across patients and test sessions from the sample of 32 odours. For the discrimination test, the order of triplets was also randomised across patients and test sessions. The regular Sniffin’ Sticks test battery was used for the control group, and all controls performed the same presentation order in the discrimination and identification tasks.

Results of the three subtests are presented as a composite “Sniffin’ Sticks test score” (TDI score, range 1-48), which is the sum of the results obtained for threshold, discrimination and identification measures, with higher scores indicating better olfactory function. A Sniffin’ Sticks test score of 16.25 and below is defined as anosmia; a score between 16.5 and 30.5 as hyposmia; and a score of 30.75 and above as normosmia [40].

For the patients in the home visit cohort, the Sniffin’ Sticks test score of the home visit preceding the MRI scan was used in the current analysis.

#### 2.2.3. Assessment of parosmia symptoms in patients

The presence of parosmia symptoms in patients was assessed through an online questionnaire. Similar to Parma et al. [9], patients were asked whether they experienced any of the following symptoms in a check-all-that-apply question: (1) “I cannot smell at all”; (2) “Odours smell less strong than they did before”; (3) “Odours smell different than they did before (the quality of the odour has changed)”; (4) “I can smell things that are not there (for example, I smell fire when nothing is on fire)”; (5) “Sense of smell fluctuates (comes and goes)”; and (6) “None of the above”. Option 3 was used to classify patients as parosmic.

For patients in the longitudinal COVORTS study, this assessment was performed at home (see van Dijk et al. [26] for more information on the cohort design). For patients who were part of the MRI sub-study, this questionnaire was completed during the screening session in which the Sniffin’ Sticks test battery was administered. For patients in the home visit cohort, data from the questionnaire for the home visit preceding the MRI scan were used in the current analysis. If patients were hyposmic or normosmic according to their Sniffin’ Sticks test scores and reported parosmia, they were classified as parosmic. If patients reported parosmia but scored within the anosmia criteria, they were classified as having quantitative OD.

#### 2.2.4. MRI data acquisition

Scans were acquired on a 3-Tesla MRI scanner (Elition X, Philips Medical Systems, Amsterdam, The Netherlands) using a 32-channel head coil. To image the olfactory bulbs a coronal T*_2_*-weighted scan was made (repetition time (TR): 3152 ms, echo time (TE): 165 ms, field of view (FOV): 120 × 120 x 28 mm, acquisition of 28 coronal slices, acquired voxel size: 0.5 x 0.61 x 1.0 mm, reconstructed voxel size: 0.34 x 0.34 x 1.0 mm, and flip angle = 90°. To obtain grey matter volumes, a high-resolution T*_1_*-weighted 3D TFE whole brain anatomical scan was conducted with the following parameters: TR: 10 ms, TE: 4.6 ms, FOV: 256 x 243 x 180 mm, acquisition of 450 sagittal slices, acquired voxel size: 0.8 x 0.8 x 0.8 mm, reconstructed voxel size: 0.4 x 0.4 x 0.4 mm, and flip angle: 8°. Finally, functional scans were conducted using a T*_2_\**-weighted gradient echo 2D-EPI sequence, with the following parameters: TR: 0.95 ms, TE: 25 ms, SENSE factor (AP): 1.7, multiband factor: 4, FOV: 208 x 208 x 132 mm, acquisition of 60 axial slices in ascending order, acquired voxel size: 2.2 x 2.2 x 2.2 mm, reconstructed voxel size: 2.17 x 2.17 x 2.2 mm, flip angle: 57°.

#### 2.2.5. fMRI task

The fMRI paradigm and odours used in this study are the same as described by Reichert et al. [44]. Olfactory stimulation during the functional scan was performed using an 8-channel computer-controlled olfactometer (Burghart Messtechnik GmbH, Holm, Germany). Odours were administered orthonasally to the patient using two nosepieces inserted into the nostrils. Two odour stimuli equivalent in intensity were used: chocolate (IFF, SC048015; 8.5% dissolved in propylene glycol) and beef (IFF, 10920656; 0.04% dissolved in demineralised water). Odour stimuli were embedded in a stream of odourless, humidified air (80%, air flow 8 L/min, 36 °C). Additionally, blank trials were incorporated, during which the visual cue indicating odour stimulation was presented for the same duration as the odour trial, whilst no actual odour was administered. Presentation of visual cues and pictures, and triggering of the olfactometer, were performed using E-Prime 3.0 (Psychology Software Tools, Inc.).

The fMRI task (see Fig. 1) consisted of two 15-minute blocks separated by a one-minute break. Before the actual tasks started, a set of warm-up questions was presented to familiarise participants with answering questions on the visual analogue scales (VAS). Patients responded to these questions by moving a cursor along the VAS line by pressing buttons on a button box. After the test questions, participants were instructed that the task would start and were shown a white fixation cross on a black screen. Participants were told they could breathe through either their nose or mouth during the white fixation cross, and to breathe through their nose once the fixation cross turned red. After the first block was completed, participants saw a screen indicating that they had a one-minute break before the second block would start.

In total, 20 chocolate odour trials, 20 beef trials and 20 blank trials were presented. In addition, ten combined chocolate & picture and ten beef & picture trials were presented. During these combined odour-picture trials, patients were shown a picture of a chocolate muffin or a steak in addition to the odour administration. All trials were preceded by the white fixation cross turning red. Odour release occurred 1000 ms after the first appearance of the red cross and lasted 2000 ms. In the odour-picture trials, the red cross visual was replaced with the food picture 200 ms after odour release. In the non-picture trials, the red cross remained on screen for another 2000 ms (2200 ms after odour release, 3200 ms since first appearance). To sustain participants’ attention, participants answered questions on a VAS (range 0-100). The cursor always started in the centre of the VAS. Half of the odour/blank trials were followed by the question “How intense did you perceive the odour?” with the anchors ‘not strong at all” and “very strong”. Eleven of the combined odour & picture trials were followed by the question “How well did the picture and the odour match?” with the anchors “not matching at all” and “very matching”. The inter-trial interval ranged from 11 to 20 s.

**Fig. 1.**
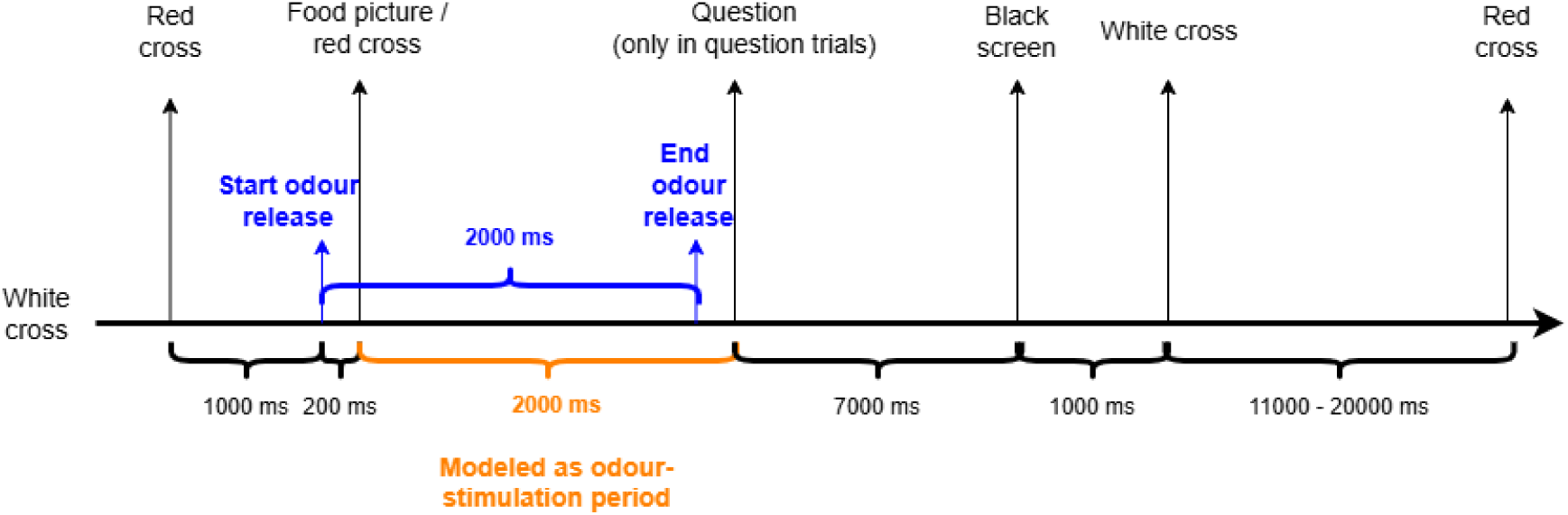
Details on stimulus timing during the olfactory paradigm (not to scale). The blue interval is the period of odour release. The orange interval represents the duration modelled as odour stimulation in the analysis.

### 2.3. Data analysis

Unless stated otherwise, statistical analyses were conducted in R (version 4.4.2). For the patients, three groups were made for the analyses: (1) grouped OD, including all patients, (2) quantitative OD, including patients with a Sniffin’ Sticks test score ranging within anosmia and hyposmia (with no reported parosmia symptoms), and (3) parosmia, including all patients who reported parosmia symptoms (with a Sniffin’ Sticks test score ranging within hyposmia and normosmia). For most analyses, these three groups were each compared to the normosmic controls. Furthermore, the patients with quantitative OD and those with parosmia were also compared to each other. Demographic variables were compared using two-sample *t*-tests and Analysis of Covariance (ANCOVA).

#### 2.3.1. Olfactory bulb volume

Manual segmentation of the olfactory bulbs was performed using ITK-SNAP (version 4.0.2, University of Pennsylvania & University of Utah, www.itksnap.org) [45]. The sudden change in diameter at the beginning of the olfactory tract was used as the boundary of the olfactory bulb [46]. Volumes were obtained by planimetric manual contouring of the surface area of the bulb on each slice, resulting in a volume in cubic millimetres [32, 47]. Manual segmentation was performed by two trained observers independently (PS and BD). When the two measurements of a particular participant differed by more than 10%, the observers compared their segmentations to reach consensus. In two cases in the patient group, no visible OBs were present. A third trained observer (EP) was consulted to confirm these findings. These two cases involved one female with parosmia and one male with quantitative OD (Sniffin’ Sticks test score result within the range of anosmia). These two patients were not included in the further OBV analysis. For the remaining participants, the average of the two manual segmentation measurements was used in the statistical analysis.

To evaluate differences in OBV between groups, two-sample *t*-tests were used. Additionally, ANCOVA models were employed to test for differences between groups correcting for age, sex, and total intracranial volume (TIV). Correlation analysis between OBV and Sniffin’ Sticks test score and number of days since SARS-CoV-2 diagnosis was performed using Spearman’s rank correlations.

#### 2.3.2. Grey matter density

Voxel-based morphometry (VBM) analysis was performed using the CAT12 toolbox (vCAT12.9) [48] implemented within the SPM12 software package (v7771, Wellcome Centre for Human Neuroimaging, UCL, London, UK, http://www.fil.ion.ucl.ac.uk/spm) in MATLAB R2021a (The Mathworks, Inc., Natick, MA, USA). Using the default settings in SPM12, the whole-brain *T_1_-weighted* scans were segmented into grey matter, white matter and cerebrospinal fluid. The volumes of these segmentations, including total intracranial volume (TIV), were extracted. Subsequently, the GM segmentations were normalised and smoothed using a Gaussian kernel (full width at half maximum of 6 mm).

Differences in GMD between patients and controls were investigated using whole-brain and region-of-interest (ROI) analyses. An ROI mask was created using the primary olfactory cortex and the secondary olfactory-related regions from the merged structural and functional network map for olfactory neuroimaging, as proposed by Fjaelstad et al. [49]. The Automated Anatomical Labelling (AAL) atlas [50] in the WFU pickatlas toolbox [51] was used to create an ROI mask containing the following regions (bilaterally): Olfactory, Frontal_Sup_Orb, Frontal_Mid_Orb, Frontal_Med_Orb, Frontal_Inf_Orb, Olfactory, Rectus, Insula, Cingulum_Ant, Hippocampus, ParaHippocampal, Amygdala, Caudate, Putamen, Temporal_Pole_Sup and Temporal_Pole_Mid.

Differences in GMD between patients and controls were examined using two-sample *t*-tests, with age, sex, and TIV included as covariates. One-sample *t*-tests including Sniffin’ Sticks test score and the number of days since SARS-CoV-2 diagnosis were used to test for correlations between these variables and GMD in the grouped OD patients (also correcting for age, sex and TIV, as well as additional models correcting for Sniffin’ Sticks test score). To ensure that only grey matter regions were included in the statistical analysis, we applied an absolute masking threshold of 0.1. For the whole-brain analysis, a cluster-level threshold of *p* < 0.05, family-wise error (FWE)- corrected, was applied in conjunction with a cluster extent threshold of k > 20 voxels. This yielded no significant results. ROI analysis using a cluster-forming threshold of *p*=0.001 (uncorrected) also yielded no significant results at a cluster-level *p*_fwe_ < 0.05. Therefore, we performed exploratory ROI analyses using a cluster-forming threshold of *p*=0.001 (uncorrected) in conjunction with a cluster extent threshold equal to the expected number of voxels per cluster for each respective model [48]. Any cluster with a cluster size above this extent threshold was considered significant. Using the Diffeomorphic Anatomical Registration through Exponentiated Lie algebra (DARTEL) toolbox, grey matter segmentations from all participants were used to create a group-specific template for presenting results. Clusters were identified using the AAL3 brain atlas [52] in MRIcron (v1.0.20190902) and Neuromorphometrics in SPM when a more precise position description was needed (e.g. anterior, posterior). Mean parameter estimates were extracted from clusters using the MarsBar SPM toolbox [53].

#### 2.3.3. Odour-induced brain activation

Functional data were pre-processed and analysed using SPM12 run within MATLAB R2021a. First, functional images were realigned, and slice-time correction was performed. Anatomical images were co-registered to the mean functional image and normalised into MNI space. After this, functional images underwent smoothing, using a 3D isotropic Gaussian kernel (full width at half-maximum 4.34 mm). Finally, the ArtRepair toolbox was used to reduce the residual errors of more than 0.5 mm movement between scans which remained after realignment [54]. With the exception of one patient and two normosmic controls (22.4%, 22.8% and 25.3%, respectively), the percentage of repaired volumes was below 20% (range: 0.1 – 18.6%). Nevertheless, no participants were excluded from analysis at this step.

##### 2.3.3.1. Subject-level analysis

The following conditions were modelled: chocolate odour, beef odour, blanks, chocolate odour with a congruent picture, and beef odour with a congruent picture. The duration of the odour stimulation trials was 2000 ms, and the exact timing of the modelled stimulus is shown in Fig. 2. The six motion regressors were included to account for motion-related variance. For each subject, contrasts were made comparing all odour trials (chocolate and beef odour, plus the trials with the food picture), henceforth called “OdourTot” versus the “blank” and “rest” conditions, as well as the separate trials with only the odours (OdourOnly) and the trials with only the odour accompanied by the food picture (OdourPic). The OdourPic-versus-blanks contrast was used to assess visual cortex responses as an additional quality check. Two patients (one male patient with quantitative OD, one female patient with parosmia) and two control participants (both females) showed very little to no activation in the visual cortex and were excluded from the group-level analysis. Finally, one control did not complete the fMRI scan during the session, resulting in a sample of 46 patients and 46 controls in the group-level analysis.

##### 2.3.3.2. Group-level analysis

Two-sample *t*-tests were used to compare patients and controls using the ‘OdourTot versus rest’ and ‘OdourTot versus blanks’ contrast images. In all analyses, age and sex were added as covariates. Whole-brain analysis was performed using a cluster-level threshold of *p* < 0.05, family-wise error (FWE)- corrected, in conjunction with a cluster extent threshold of k=20 voxels, which yielded no significant results. ROI analysis, using the same mask as described in section 2.3.2, was performed with a cluster-forming threshold of *p*=0.001 (uncorrected) and yielded no significant results at cluster-level *p*_fwe_ < 0.05. Furthermore, one-sample *t*-tests were conducted on the Sniffin’ Sticks test score and the number of days since SARS-CoV-2 diagnosis to assess correlations between these variables and brain activation in the grouped OD patients. This also yielded no significant results. Therefore, one-sample *t*-tests were performed for timepoint to evaluate activation patterns, using a cluster-forming threshold of *p*=0.001 (uncorrected). Clusters were regarded as significant when (cluster-level) *p*_fwe_ < 0.05. Significant clusters were identified using the AAL3 brain atlas [52] in MRIcron (v1.0.20190902) and Neuromorphometrics in SPM when a more precise position description was needed (e.g. anterior, posterior). Mean anatomical images of all participants, as well as for each respective analysis group, were calculated to present results on, and mean parameter estimates were extracted from the clusters using the MarsBar SPM toolbox [53].

## 3. Results

### 3.1. Participant characteristics

Table 1 summarises the demographic and clinical characteristics of the study participants. The study sample included 49 normosmic controls and 48 patients with OD, of whom 21 were classified as quantitative OD (8 anosmic and 13 hyposmic) based on their Sniffin’ Sticks test score, and 27 had self-reported parosmia (qualitative) complaints (24 hyposmic and 3 normosmic). Participants in the control group were younger than those in the patient group and had an even sex distribution, whereas the patient group mainly consisted of women (79%).

**Table 1.** Participant demographics and clinical characteristics.

|  | Controls<br>(n=49) | Grouped OD<br>(n=48) | Quantitative<br>OD (n=21) | Parosmia<br>(n=27) |
| --- | --- | --- | --- | --- |
| Patient subgroup |  |  |  |  |
| Age, mean ± SD (range) | 39.9 ± 13.5<br>(22-60) | 47.4 ± 9.5<br>(19-60) | 44.7 ± 11.3<br>(19-58) | 49.5 ± 7.5<br>(31-60) |
| Sex, nr females/males | 25/24 | 38/10 | 15/6 | 23/4 |
| Sniffin' Sticks score, mean ± SD<br>(range) | 34.7 ± 2.7<br>(31-41.5) | 22.6 ± 6.8<br>(6-33.25) | 20.1 ± 8.2<br>(6-29.5) | 24.6 ± 4.7<br>(16.75-33.25) |
| Days since SARS-CoV-2 diagnosis,<br>mean ± SD (range) | N/A | 472 ± 472<br>(86-1500) | 591 ± 549<br>(90-1500) | 379 ± 387 (86-1221) |
| Severity of infection |  |  |  |  |
| *Non-symptomatic/mild* | N/A | 20.8% | 23.8% | 18.5% |
| *Moderate* | N/A | 66.7% | 52.4% | 77.8% |
| *Severe (hospital admission)* | N/A | 12.5% | 23.8% | 3.7% |
| *Critical (intensive care admission)* | N/A | 0% | 0% | 0% |
| Total intracranial volume (ml),<br>mean ± SD (range) | 1472 ± 157 | 1430 ± 147 | 1430 ± 153 | 1430 ± 145 |
| Total grey matter volume (ml),<br>mean ± SD (range) | 686 ± 81 | 645 ± 67 | 644 ± 71 | 645 ± 64 |
| Total white matter volume (mL),<br>mean ± SD (range) | 518 ± 73 | 508 ± 62 | 504 ± 61 | 511 ± 63 |

Patients tested positive for SARS-CoV-2 between April 2020 and February 2023. The time between infection and inclusion ranged from three months to over four years. Most patients had mild to moderate symptoms during the acute infection, and all but one of the severe cases were patients with quantitative OD.

The patient groups, separated into quantitative OD and parosmia and grouped together, scored significantly lower on the Sniffin’ Sticks test than the normosmic controls (all comparisons *p* < 0.001). Patients with parosmia scored slightly higher on olfactory function than patients with quantitative OD (*p* = 0.035). There were no significant differences in total intracranial volume (TIV) or total white matter volume between groups. Patients, both grouped and separated into their respective OD groups, had lower total grey matter volume compared with controls (*p* = 0.006 for grouped OD, *p*=0.034 for patients with quantitative OD, and *p*=0.017 for patients with parosmia). All of these comparisons remained significant after adjusting for age and sex: *p* < 0.001, *p*=0.002, and *p*=0.001, respectively.

### 3.2. Olfactory bulb volume

Demographics of participants in the OBV analysis (i.e., excluding the two patients without visible OBs) are provided in Appendix A, Supplementary Table S1. Olfactory bulb volumes for the different groups are reported in Table 2. When grouped, OD patients had a significantly lower total OBV compared to controls (mean difference = −10.9, 95% CI [-20.4, −1.3], t(92) = −2.26, *p*=0.026). Both patient groups showed a marginally significantly smaller OBV than the controls (patients with quantitative OD: mean difference = −11.1, 95% CI [-23.5, 1.4], t(40) = −1.80, *p*=0.079; patients with parosmia: mean difference = −10.7, 95% CI [-21.8, 0.4], t(58) = −1.92, *p*=0.058). There was no significant difference in total OBV between the two patient groups (*p*=0.957). Lastly, controlling for age, sex, and total intracranial volume did not meaningfully alter this pattern of results (grouped OD versus controls: *p*=0.027; quantitative OD versus controls: *p*=0.091; parosmia versus controls: *p*=0.072; and quantitative OD versus parosmia: *p*=0.985).

**Table 2.** Mean ± SD olfactory bulb volume (mm^3^) of patients with persistent COVID-19-related OD and normosmic controls.

|  | Normosmic controls<br>(n=49) | Grouped OD<br>(n=46) | Quantitative OD<br>(n=20) | Parosmia (n=26) |
| --- | --- | --- | --- | --- |
| Left OBV | 48.5 ± 13.7 | 43.4 ± 12.7 | 43.3 ± 13.3 | 43.5 ± 12.3 |
| Right OBV | 46.8 ± 13.0 | 41.0 ± 9.4 | 40.9 ± 9.3 | 40.9 ± 9.3 |
| Total OBV | 95.3 ± 25.3 | 84.4 ± 21.4 | 84.2 ± 22.0 | 84.6 ± 21.2 |

There was a weak positive correlation between OBV and Sniffin’ Sticks test score for all patients and controls grouped together (n=95, *r*_s_=0.26, *p*=0.011). There was a marginally significant, weak positive correlation between OBV and Sniffin’ Sticks test score when all patients were grouped together (*r*_s_=0.28, *p*=0.060). However, when examining the groups separately, there were no significant correlations between OBV and quantitative olfactory functioning (*p*=0.634 for normosmic controls, *p*=0.213 for patients with quantitative OD, and *p*=0.237 for patients with parosmia). Finally, there was no significant correlation between OBV and the number of days since SARS-CoV-2 diagnosis (*p*=0.598 for grouped OD, *p*=0.587 for patients with quantitative OD and *p*=0.164 for patients with parosmia).

### 3.3. Grey matter density

Both whole-brain and ROI analyses at the corrected threshold level yielded no significant results. The results below are therefore from the exploratory analyses.

As an additional exploration, all GMD analyses were performed with the two patients without visible OBs omitted from the sample. Overall, the results were similar to those of the full patient sample presented below. The results for this adjusted patient sample are presented in Appendix B, Supplementary Tables S2–S6.

#### 3.3.1. Grouped OD patients versus controls

The comparison between the grouped OD patients and normosmic controls revealed that patients have lower GMD in seven clusters (Table 3 and Fig. 3).

**Table 3.** Brain regions with lower grey matter density for grouped COVID-19 OD patients (n=48) compared to normosmic controls (n=49) within ROIs (exploratory analysis*).

| Brain region | Cluster size (voxels) | Hemisphere | Peak voxel MNI coordinate |  |  | Z-score |
| --- | --- | --- | --- | --- | --- | --- |
|  |  |  | X | Y | Z |  |
| Anterior orbital gyrus<sup>1</sup> | 74 | R | 17 | 59 | -14 | 4.61 |
| Middle frontal gyrus<sup>2</sup> | 84 | R | 45 | 53 | -12 | 4.04 |
| Anterior orbital gyrus<sup>3</sup> | 406 | L | -20 | 59 | -14 | 4.00 |
| Medial orbital gyrus |  | L | -9 | 54 | -23 | 3.76 |
| Medial orbital gyrus |  | L | -11 | 42 | -20 | 3.63 |
| Nucleus accumbens<sup>4</sup> | 81 | R | 11 | 12 | -12 | 3.85 |
| Caudate nucleus<sup>5</sup> | 63 | R | 6 | 12 | 0 | 3.73 |
| Medial orbital gyrus | 64 | R | 14 | 38 | -20 | 3.71 |
| Gyrus rectus |  | R | 6 | 32 | -27 | 3.14 |
| Olfactory cortex<sup>6</sup> | 65 | L | -14 | 12 | -14 | 3.69 |

\*Results presented are clusters with an extent threshold equal to the expected number of voxels per cluster; k=44 at a cluster-forming threshold of $p=0.001$ (uncorrected).
<sup>1</sup>Cluster extends into the right dorsolateral and medial orbital superior frontal gyrus.
<sup>2</sup>Cluster extends into the right anterior OFC and IFG pars orbitalis.
<sup>3</sup>Cluster extends into the left dorsolateral superior frontal gyrus, gyrus rectus, and middle frontal gyrus.
<sup>4</sup>Cluster extends into the right gyrus rectus and olfactory cortex.
<sup>5</sup>Cluster extends into the right nucleus accumbens.
<sup>6</sup>Cluster extends into the left gyrus rectus, nucleus accumbens and putamen.

**Fig. 3.**
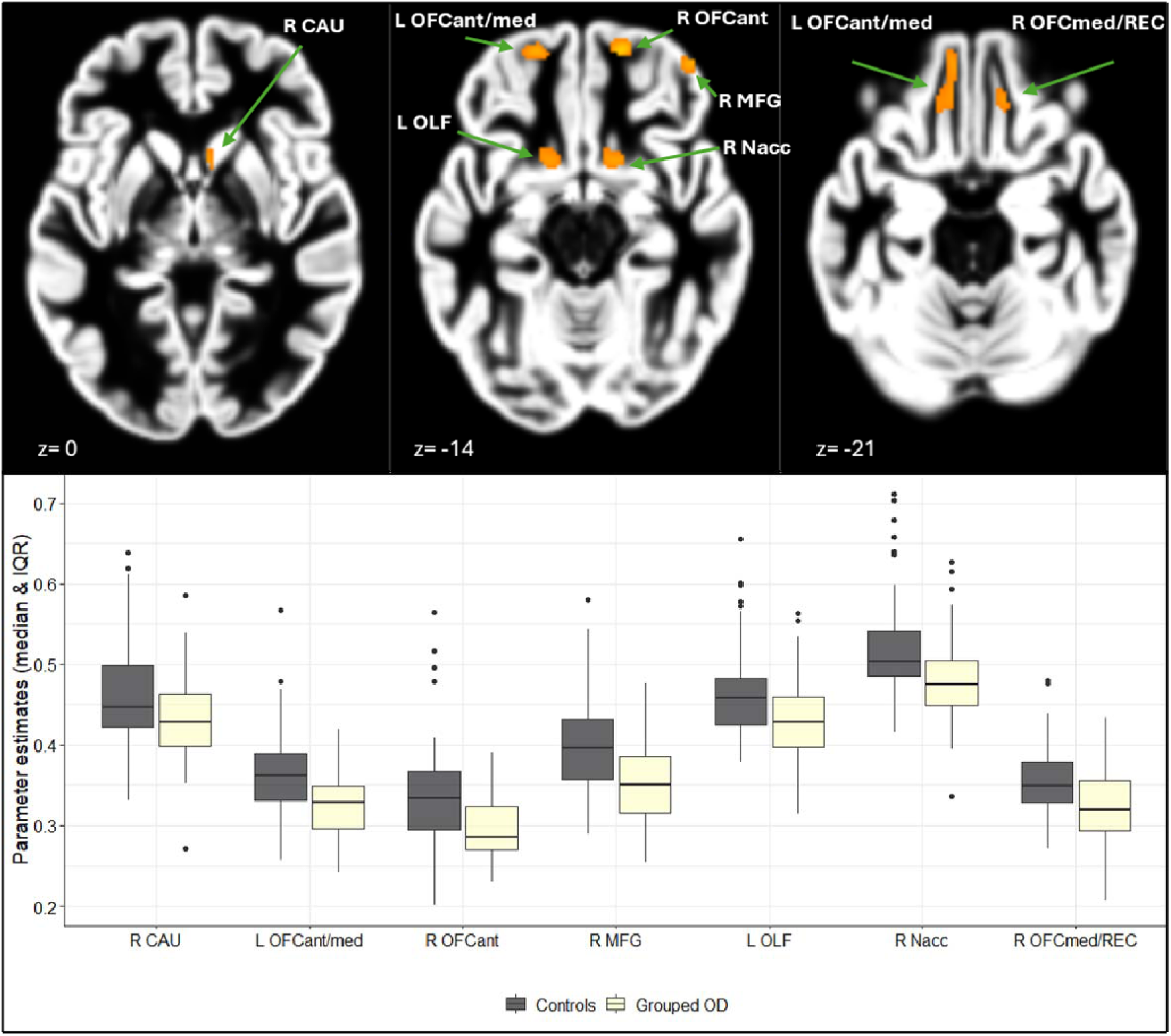
Colour-coded T-maps thresholded at T=3.18 (cluster-forming threshold *p*=0.001 (uncorrected), cluster extent threshold k=44 (expected number of voxels)) showing lower GMD in grouped COVID-19 OD patients (n=48) versus normosmic controls (n=49) within ROIs, overlaid onto the mean sample GM image (top) and boxplot of associated average parameter estimates (bottom). CAU = caudate; OFCant = anterior orbital gyrus; OFCmed = medial orbital gyrus; MFG = middle frontal gyrus; OLF = olfactory cortex; Nacc = nucleus accumbens; REC = gyrus rectus. The lower and upper whiskers of the boxplots represent the smallest and largest values within 1.5 times the interquartile range (IQR), respectively. Data points beyond 1.5 times the interquartile range are plotted individually.

#### 3.3.2. Patients with quantitative OD versus controls

The comparison between patients with quantitative OD and normosmic controls showed lower grey matter density in patients in eight clusters (Table 4 and Fig. 4).

**Table 4.** Brain regions with lower grey matter density for COVID-19 OD patients with quantitative OD (n=21) compared to normosmic controls (n=49) within ROIs (exploratory analysis*).

| Brain region | Cluster size (voxels) | Hemisphere | Peak voxel MNI coordinate |  |  | Z-score |
| --- | --- | --- | --- | --- | --- | --- |
|  |  |  | X | Y | Z |  |
| Anterior cingulate cortex, pregenual<sup>1,**</sup> | 96 | R | 2 | 54 | 11 | 5.22 |
| Superior frontal gyrus, medial |  | L | 3 | 50 | 18 | 3.62 |
| Medial orbital gyrus | 81 | R | 14 | 39 | -20 | 4.06 |
| Gyrus rectus |  | R | 6 | 33 | -27 | 3.43 |
| Medial orbital gyrus<sup>2</sup> | 189 | L | -11 | 42 | -20 | 3.81 |
| Medial orbital gyrus |  | L | -9 | 56 | -23 | 3.76 |
| Medial orbital gyrus |  | L | -14 | 29 | -27 | 3.59 |
| Anterior orbital gyrus<sup>3</sup> | 64 | R | 44 | 53 | -14 | 3.79 |
| Nucleus accumbens<sup>4</sup> | 59 | R | 12 | 11 | -12 | 3.78 |
| Superior frontal gyrus, dorsolateral | 102 | L | -23 | 66 | -6 | 3.74 |
| Superior frontal gyrus, dorsolateral |  | L | -12 | 66 | -12 | 3.39 |
| Nucleus accumbens<sup>5</sup> | 114 | L | -5 | 8 | -6 | 3.55 |
| Olfactory cortex |  | L | -14 | 11 | -14 | 3.52 |
| Caudate nucleus | 43 | R | 11 | 17 | 11 | 3.25 |
| Caudate nucleus |  | R | 6 | 17 | 3 | 3.11 |

\*Results presented are clusters with an extent threshold equal to the expected number of voxels per cluster; k=42 at a cluster-forming threshold of $p=0.001$ (uncorrected).
\*\*Peak voxel significant at $p_{\text{fwe}} < 0.05$ .
<sup>1</sup>Cluster extends into the left subgenual anterior cingulate cortex and the right medial superior frontal gyrus.
<sup>2</sup>Cluster extends into the left gyrus rectus.
<sup>3</sup>Cluster extends into the right IFG pars orbitalis and middle frontal gyrus.
<sup>4</sup>Cluster extends into the right gyrus rectus and olfactory cortex.
<sup>5</sup>Cluster extends into the left gyrus rectus and putamen.

**Fig. 4.**
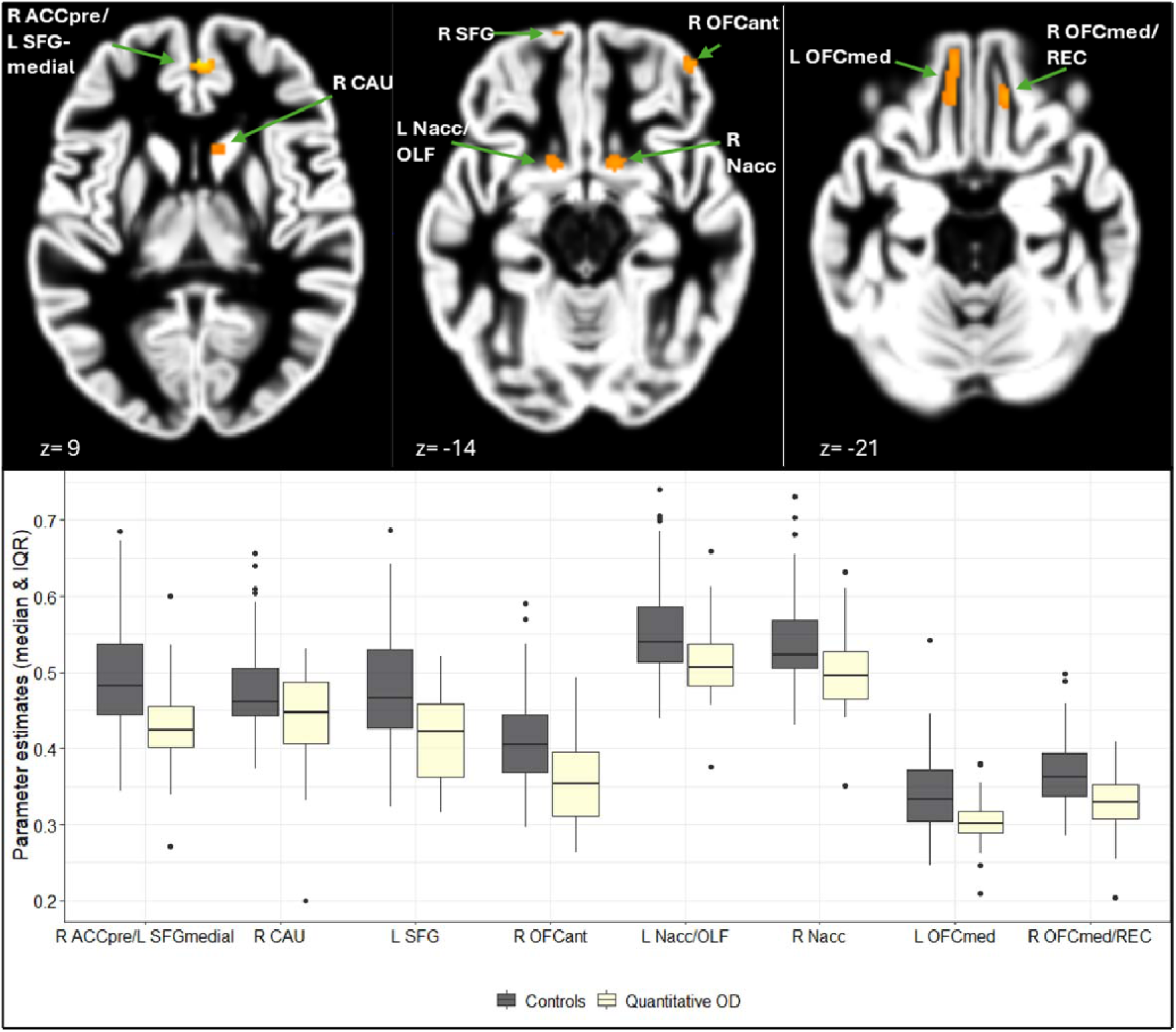
Colour-coded T-maps thresholded at T=3.18 (cluster-forming threshold *p*=0.001 (uncorrected), cluster extent threshold k=42 (expected number of voxels)) showing lower GMD in COVID-19 OD patients with quantitative OD (n=21) versus normosmic controls (n=49) within predefined ROIs, overlaid onto the mean sample GM image (top) and boxplot of associated average parameter estimates (bottom). ACCpre = anterior cingulate cortex, pregenual; SFGmedial = superior frontal gyrus, medial; CAU = caudate; SFG = superior frontal gyrus, dorsolateral; OFCant = anterior orbital gyrus; Nacc = nucleus accumbens; OLF = olfactory cortex; OFCmed = medial orbital gyrus; REC = gyrus rectus. The lower and upper whiskers of the boxplots represent the smallest and largest values within 1.5 times the interquartile range (IQR), respectively. Data points beyond 1.5 times the interquartile range are plotted individually.

#### 3.3.3. Patients with parosmia versus controls

The comparison between patients with parosmia (n=27) and normosmic controls (n=49) revealed that patients show lower GMD in the left anterior orbital gyrus (k=80, Z=3.72, MNI (−21, 59, −15), expected number of voxels per cluster k=41), with the cluster extending into the middle and superior frontal gyrus (dorsolateral) (see Appendix B, Supplementary Fig. S1). However, this cluster no longer appeared when controlling for Sniffin’ Sticks test score in the model.

#### 3.3.4. Patients with quantitative OD versus patients with parosmia

There was no difference in GMD between the two patient groups at the exploratory threshold.

#### 3.3.5. Correlations with Sniffin’ Sticks test score and days since SARS-CoV-2 infection

Within the grouped OD patients (n=48), there was a positive correlation between GMD and Sniffin’ Sticks test score at a cluster in the left olfactory cortex, extending into the left nucleus accumbens (k=49, Z=3.91, MNI (−11, 11, −14), expected number of voxels per cluster k=35). No correlation was found between the number of days since SARS-CoV-2 diagnosis and GMD within the patient group.

### 3.4. Odour-induced brain activation

Demographics of the sample used in the fMRI analysis are presented in Appendix C, Supplementary Table S7.

Both the whole-brain and ROI analyses showed no difference in odour-induced brain activation between patients and controls (OdourTot vs rest/blanks). This did not change when excluding the two patients without visible OBs. No correlations were found between Sniffin’ Sticks test score and odour-induced brain activation, nor for the number of days since SARS-CoV-2 diagnosis.

Significantly activated ROIs for each group in response to odour stimulation (vs rest) are presented in Fig. 5 and Appendix C, Supplementary Tables S8-S11, and Supplementary Figs. S2-S5. All groups generally showed activation in the same five clusters areas: anterior cingulate cortex (supracallosal), bilateral anterior insula / IFG pars orbitalis (extending into the olfactory cortex, putamen, gyrus rectus, temporal pole: superior and middle temporal gyrus, medial and posterior orbital gyri, caudate nucleus, amygdala, nucleus accumbens, middle frontal gyrus, dorsolateral superior frontal gyrus, Rolandic operculum, hippocampus and parahippocampal gyrus); and bilateral hippocampus / parahippocampal gyrus / lateral geniculate/lingual gyrus.

**Fig. 5.**
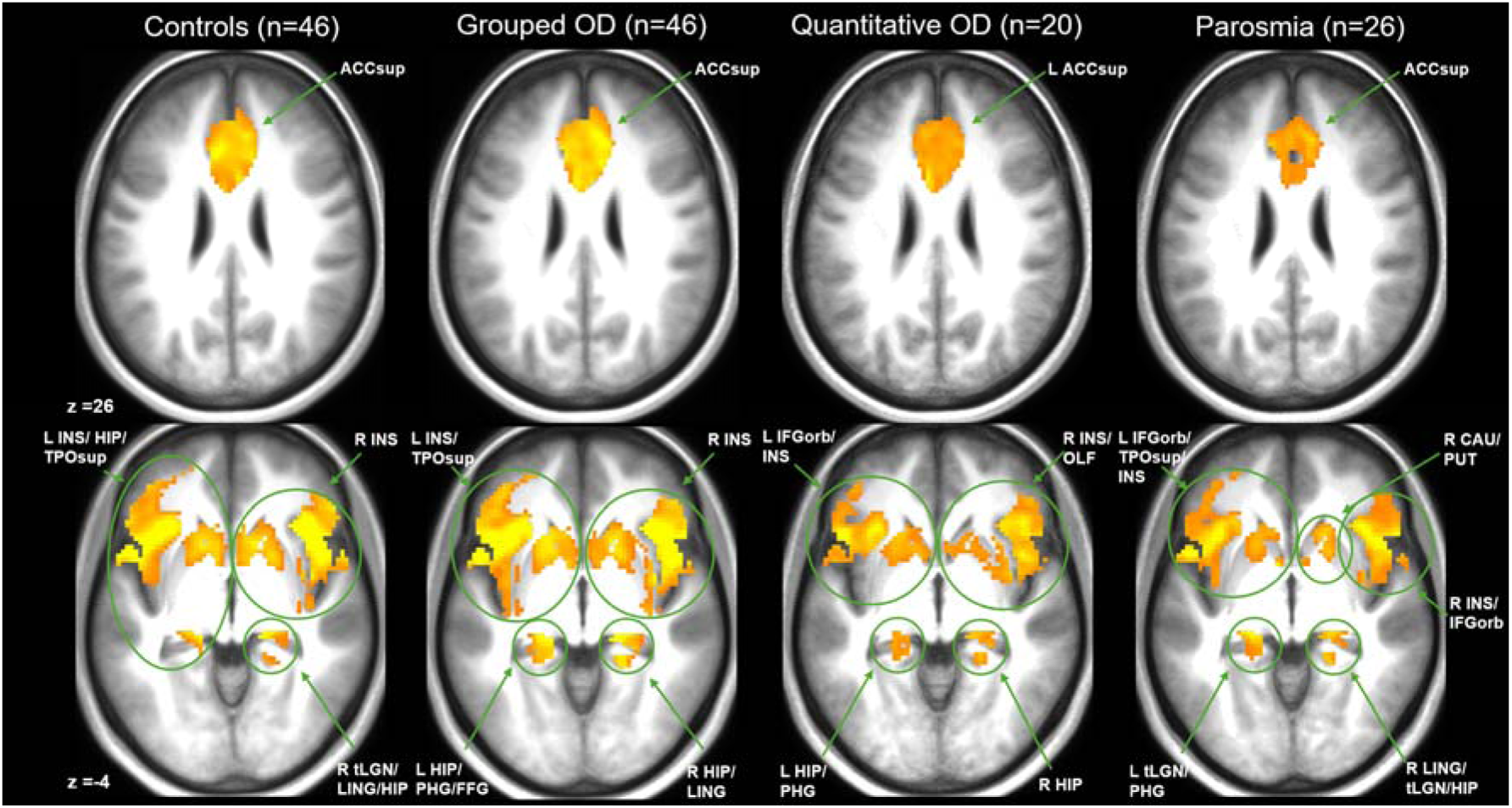
Colour-coded T-maps thresholded at T=3.29 (cluster-forming threshold *p*=0.001 (uncorrected)), showing odour-induced brain activation (OdourTot > rest contrast) per group, overlaid onto the respective mean anatomical image. Shown clusters are significant at cluster-level *p*_fwe_ < 0.05. ACCsup = anterior cingulate cortex, supracallosal; INS = insula (anterior); HIP = hippocampus; TPOsup = temporal pole: superior temporal gyrus; tLGN = lateral geniculate; LING = lingual gyrus; PHG = parahippocampal gyrus; FFG = fusiform gyrus; IFGorb = IFG pars orbitalis; OLF = olfactory cortex; CAU = caudate; PUT = putamen.

Regarding odour stimulation versus the blank trials, the controls, grouped patients, and patients with parosmia showed activation in the left parahippocampal gyrus, hippocampus (bilateral), and amygdala (bilateral) (Appendix C, Supplementary Tables S8-S11, and Supplementary Fig. S6).

## 4. Discussion

This comprehensive (f)MRI study examined the neural correlates of persistent COVID-19-related quantitative and qualitative olfactory dysfunction (OD). Both structural and functional neural alterations were investigated by examining olfactory bulb volume (OBV), grey matter density (GMD) and brain activation in response to odour stimuli. This was performed in patients with persistent OD, separated into a group of patients with quantitative OD and a group of patients with parosmia symptoms, as well as a group of normosmic controls. Patients with persistent COVID-19-related olfactory dysfunction (C19OD) showed smaller OBV compared to controls and a tendency towards lower GMD in primary and secondary olfactory cortices. However, analysis of odour-induced brain activation (BOLD-response) showed no differences between patients and controls, with all groups exhibiting a similar activation pattern.

### 4.1. Olfactory bulb volume

The smaller OBV in patients with persistent C19OD in comparison with healthy controls is in line with previous studies [31, 33, 55–57]. Our total sample size is similar to that of these previous studies. Still, when we classified our patients as having quantitative OD or parosmia, the differences were no longer statistically significant.

We did not find a difference in OBV between patients with quantitative OD and those with parosmia. Pre-SARS-CoV-2 pandemic studies have shown that patients who suffer from co-occurring parosmia and quantitative OD presented with smaller OBV than those with only quantitative OD [58, 59]. This discrepancy may be due to the small sample sizes of our subgroups or to our diagnostic criteria for parosmia. Although quantitative OD and parosmia often co-occur in post-COVID and pre-pandemic cases of OD [60], we diagnosed any patient who scored within the anosmic range on the Sniffin’ Sticks test as having quantitative OD. This decision was made because we presumed that any patient with no functional sense of smell (i.e., anosmia) could not experience parosmia symptoms, and therefore misinterpreted our question measuring the presence of parosmia. However, patients with parosmia may potentially score lower on the Sniffin’ Sticks test due to their distorted odour perception, making the discrimination and identification subtests more difficult. This diagnostic criterion may have limited the accuracy of our diagnosis of anosmia and parosmia.

Although we observed a minimal correlation between OBV and Sniffin’ Sticks test score, pre-pandemic reports have found a positive association between volume of the olfactory bulbs and quantitative olfactory functioning [61, 62]. Nevertheless, as previous COVID-19 OD studies often did not include information on the presence of parosmia symptoms [31, 33, 55–57], our study is one of the first to show that – whilst not different from each other – both patients with COVID-19-related quantitative OD and parosmia tend to have smaller OBV than normosmic controls.

The two patients without visible OBs were a striking finding of the current investigation. Altered morphology in the form of deformed shapes of the OBs has been reported in other studies on C19OD [29, 30], but only one report mentions the absence of OBs. In their investigation of 36 patients with persistent C19OD, Altunisik and colleagues [31] characterised four patients as having total atrophy of the OBs. For one of these patients, they performed a follow-up 3 months later and observed no changes in the OB, but reported that her OD symptoms had improved. For these cases of total OB atrophy, one could question whether the absence of OB structures was caused by or preceded the patients’ COVID-19 infections. Notably, Weiss and colleagues [63] reported that a small percentage of women, particularly those who are left-handed, have normal olfactory function without visible OBs. We are unaware of our participants’ handedness and, obviously, lack pre-COVID MRI scans. Whereas for our parosmic female, a pre-COVID absence of OBs could be a likely, although rare, finding – especially in the context of her now suffering from C19OD – the cause of the atrophy for the male anosmic remains unclear. Nonetheless, even after excluding these two extreme cases, patients with persistent quantitative and qualitative C19OD tended to have smaller OBV than normosmic controls.

### 4.2. Grey matter density

We found that – at an exploratory threshold – patients with persistent C19OD had lower grey matter density in primary and secondary olfactory cortical areas compared to normosmic controls (including the olfactory cortex, anterior cingulate cortex, caudate nucleus, gyrus rectus, orbitofrontal cortex and putamen). One report found regional GM atrophy in patients with C19OD compared to non-COVID controls [64]. Two other reports included patients with post-acute COVID-19-related cognitive complaints and classified them into groups with and without OD [34, 36]. They found lower densities within the primary and secondary cortices in patients with OD compared to those with only cognitive complaints. However, because the sample sizes of Campabadal et al. [34] and Arrigoni et al. [64], as well as the patient subgroups in the current manuscript, are relatively small in the context of VBM analysis [65], these findings should be interpreted with caution.

Although there was no statistically significant difference in GMD between the two patient groups, the tendency towards lower GMD in patients compared to controls appears to be driven mainly by patients with quantitative symptoms rather than those with parosmia. The seven clusters showing lower GMD in the comparison between the grouped patients and controls are virtually identical to the eight clusters of lower GMD in the comparison between patients with quantitative OD and controls. Patients with parosmia showed only one cluster of lower GMD in the left OFC compared with controls, which disappeared after controlling for Sniffin’ Sticks test score. It may therefore be that the observed tendency towards lower GMD in the left OFC in patients with parosmia is mediated by their degree of co-occurring quantitative OD. However, there was also a positive correlation between Sniffin’ Sticks test score and GMD of the insula, but not the aforementioned cluster in the OFC, indicating that the degree of co-occurring olfactory loss is not the only explanation for the tendency towards lower GMD in this cluster in patients with parosmia. Furthermore, both patient groups showed lower total grey matter volume compared to controls, even when correcting for Sniffin’ Sticks test score, indicating a global effect on GMD for both patients with quantitative OD and those with parosmia. To our knowledge, there is only one pre-pandemic study specifically focussing on GMD in patients with parosmia, which reported lower regional GMD in patients with parosmia compared to healthy controls when controlling for Sniffin’ Sticks test score [66]. However, they included patients with various aetiologies, including idiopathic and post-traumatic cases of OD, which limits the possibility of comparing these findings with our post-COVID sample. All in all, our results – although modest and to be interpreted with caution given our exploratory threshold and modest sample size – suggest the possibility of grey matter degradation in patients with persistent quantitative and qualitative C19ODOD.

### 4.3. Odour-induced brain activation

We did not find differences in brain activation in response to odour stimulation between our patients and controls; all three groups exhibited a similar activation pattern. Similar task-based fMRI studies comparing post-acute COVID-19 OD patients to normosmic controls are scarce. Yildirim and colleagues [30] compared 31 patients with C19OD to 97 patients with post-viral OD using an fMRI experiment, and found no differences in brain activation. However, they focused only on activation in the OFC and entorhinal cortex, and their long single-block stimulus design (2 minutes) might yield different activation patterns than our short (2 seconds) repeated-stimulus design [67]. A pre-pandemic fMRI study has shown that people with hyposmia exhibit odour-induced activation in areas typically activated by odour stimulation, albeit to a lower degree than in healthy controls [68]. Most patients included in our analysis were hyposmic, which may have resulted in a relatively normal brain activation pattern.

On the other hand, there were some nominal differences in activation pattern between groups when comparing odour stimulation to the blank trials. Most notably, in the contrast comparing odour exposure to the blank trials, patients with quantitative OD showed no activation in response to odour stimulation. In contrast, the control group and patients with parosmia did, indicating that patients with parosmia may have increased brain activation compared to patients with olfactory loss. The blank trials are considered to more directly reflect actual odour-induced brain activation by filtering out any activation responses from conscious inhalation, as the act of sniffing is known to induce brain activation in the primary olfactory and orbitofrontal cortex [69, 70]. Unfortunately, the low number of blank trials and the small subgroup sample sizes may have resulted in underpowered comparisons. Nevertheless, these findings using the blank trials may suggest decreased activation in patients with quantitative OD compared to those with parosmia symptoms and healthy controls, although more data are needed.

### 4.4. Synthesis of neural findings

Despite the cross-sectional design of the current manuscript, which limits our ability to pinpoint the exact pathogenesis of persistent quantitative and qualitative C19OD, our findings – together with other literature investigating this topic – allow us to speculate on the possible mechanisms underlying these post-COVID symptoms.

The absence of meaningful findings beyond the OB in the current study may indicate a peripheral mechanism. The smaller OBV in the current and previous investigations of persistent C19OD may suggest a loss of sensory input from the olfactory epithelium to the OBS, which is generally thought to result in decreased volume of the OBs [19, 32]. Recent studies have reported chronic inflammatory responses in the olfactory epithelium of patients with persistent COVID-19-related hyposmia, which may prevent the epithelium from regenerating after the acute infection [71–73]. This could then lead to quantitative loss, as well as parosmia symptoms due to incomplete activation of the remaining functioning olfactory receptors [16, 17]. Ongoing inflammation in the epithelium could also explain the frequent fluctuations reported by patients in the COVORTS study, both regarding quantitative and qualitative symptoms [26, 27].

On the other hand, the observed smaller volumes could also be the result of direct injury to the OBs, as corroborated by alterations in OB signal intensities in acute [74] and persistent C19OD cases [29, 30, 75], as well as animal evidence that the viral infection and associated inflammatory processes seen in the olfactory epithelium can spread to the olfactory bulb [76, 77]. However, there are issues in interpreting signal intensities from the OB without a proper control group [78], something none of the reports on signal intensities in persistent OD had. Moreover, direct proof of persistent inflammation in the OB in human cases of persistent C19OD is currently unavailable.

Despite our speculation about a possible peripheral mechanism underlying persistent C19OD, we cannot rule out central mechanisms entirely. Top-down modulation of OBV is possible, as demonstrated in healthy subjects following lateralized olfactory training [79]. Although the results of the current manuscript are primarily limited to OBV differences, the lower total grey matter volume and the tendency towards lower regional GMD can also indicate a central cause. However, these grey matter findings may be secondary to decreased sensory signal input from the periphery [80]. Importantly, the neuroplasticity of our brains offers the potential for recovery. Still, prolonged altered sensory input could further degrade the olfactory cortices, leading to irreversible damage despite the potential resolution of the initial cause of OD [81]. We did not find a correlation between the time since SARS-CoV-2 diagnosis and our neural outcome measures. However, longitudinal studies are lacking. Due to the potential association between olfactory loss and cognitive decline [82], longer follow-up of patients – especially those with quantitative OD - is crucial to observe any further degradation or potential recovery of both olfactory function and its neural correlates.

### 4.5. Limitations

Although several limitations of the current study have already been discussed, a few other considerations should be taken into account when interpreting our findings. First of all, the Sniffin’ Sticks test used to diagnose quantitative OD, as well as the questionnaire assessing the presence of parosmia, were not performed on the same day as the scanning session. Especially for patients recruited from the longitudinal COVORTS study, for whom the olfactory outcomes from the preceding home visit were used, quite some time could have passed between measurement of olfactory function and the scanning session (up to almost three months). With the frequent fluctuations in the prevalence of quantitative and qualitative OD reported previously [26, 27], this detail of our study design could have limited our ability to properly diagnose patients with quantitative and qualitative OD.

Furthermore, at the time the data were collected from the control group (between 2022 and 2024), it was difficult to find participants without a history of COVID-19 infection. Although we screened for current normal olfactory function, we did not ask about past infections or acute loss of smell during those infections. Surprisingly, around 40% of the people we initially screened for the control group did not pass the Sniffin’ Sticks test criteria for normosmia. Several studies have found neural alterations in patients who had already recovered from a SARS-CoV-2 infection [83–85]. This begs the question whether the lack of differences seen past the OB in the current manuscript is due to both our patients and controls suffering from OB atrophy and possible further neurodegeneration. It would be interesting to perform longitudinal follow-up of those who had only acute C19OD, those with persistent symptoms, asymptomatic patients, and – if at all possible – those who were never infected to evaluate the extent of possible neural degradation or recovery following SARS-CoV-2 infection.

## 5. Conclusion

This (f)MRI study explored the neural correlates of persistent COVID-19-related olfactory dysfunction with a focus on potential differences between patients with quantitative dysfunction and parosmia symptoms. Compared with normosmic controls, patients with persistent COVID-19-related olfactory dysfunction showed lower olfactory bulb volume and a tendency toward lower grey matter density in primary and secondary olfactory-related regions, which may be mainly mediated by quantitative olfactory symptoms. No difference in brain activation in response to odour stimulation was found between patients and controls. The investigation of both structural and functional neural alterations in the same patients led to novel insights into the possible distinct neural outcomes and mechanisms underlying quantitative and qualitative COVID-19-related olfactory dysfunction. The observed smaller olfactory bulbs, along with modest findings beyond this primary olfactory structure in patients with persistent quantitative and qualitative OD, may indicate a peripheral or localised central mechanism within the olfactory bulbs. Follow-up investigation into further neural degradation or recovery is warranted to better elucidate the pathogenesis and long-term consequences of persistent COVID-19-related olfactory dysfunction.

## Acknowledgements

The authors would like to thank all patients who took part in the COVORTS study, as well as the healthy controls who participated. Special thanks to Nienke Steenstra, Eline Oerlemans, Demi Bettonvil, Maureen Coolbergen, Laudine de Bever, Carla Meijer, Catoo Krale, Femke Bootsma, Tesse van der Struijk, Amy Kroeze, Sam Kreuk, Pien Velthuijs and Saskia Markerink for their assistance in data collection and participant management. Thank you to Yanyang Huang for her assistance with using MATLAB/SPM12 to analyse the data from the fMRI paradigm. We want to acknowledge Prof. Dr. med. Thomas Hummel and Akshita Joshi, PhD (Smell and Taste Clinic, Technische Universität Dresden, Germany), for their instruction in manual segmentation of olfactory bulbs (and hospitality). The use of the 3T MRI was made possible by WUR Shared Research Facilities.

## Data availability

Upon project completion the data presented in this article will be made available: https://doi.org/10.17026/LS/YZHHK7

## Author contributions: CRediT

**Birgit van Dijk:** Conceptualization, Data curation, Formal analysis, Investigation, Methodology, Project administration, Visualization, Writing – original draft.

**Elbrich M. Postma:** Data curation, Funding acquisition, Investigation, Methodology, Project administration, Writing – review and editing.

**Louise M. Leenders:** Data curation, Investigation, Project administration, Writing – review and editing

**Puck Smits:** Formal Analysis, Investigation, Writing – review and editing.

**Digna M.A. Kamalski:** Funding acquisition, Methodology, Writing – review and editing.

**Paul A.M. Smeets:** Conceptualization, Data curation, Formal analysis, Resources, Software, Supervision, Writing – review and editing.

**Sanne Boesveldt:** Conceptualization, Funding acquisition, Methodology, Supervision, Writing – review and editing.

