## Appendices for "Neural characterisation of persistent quantitative and qualitative COVID-19-related olfactory dysfunction: The COVORTS study"

Appendix A: Supplementary materials OBV

**Table S1.** Participant demographics – OBV analysis (excluding patient with no visible OBs).*

|  | Grouped OD  (n=46) |  |  |
| --- | --- | --- | --- |
| Patient subgroup |  | Quantitative OD  (n=20) | Qualitative OD  (n=26) |
| Age, mean ± SD (range) | 47.2 ± 9.6 (19-60) | 44.2 ± 11.3 (19-58) | 49.4 ± 7.6 (31-60) |
| Gender, female/male | 37/9 | 15/5 | 22/4 |
| Sniffin’ Sticks score, mean ± SD (range) | 23.1 ± 6.4  (6-33.25) | 20.9 ± 7.8  (6-29.5) | 24.8 ± 4.6  (16.75-33.25) |
| Days since SARS-CoV-2 diagnosis, mean ± SD (range) | 444 ± 461  (86-1500) | 561 ± 545  (90-1500) | 354 ± 371  (86-1221) |
| Severity of infection |  |  |  |
| *Non-symptomatic/mild* | 22.7% | 25.0% | 19.2% |
| *Moderate* | 67.4% | 55.0% | 76.9% |
| *Severe (hospital admittance* | 10.9% | 20.0% | 3.8% |
| *Critical (intensive care admission)* | 0% | 0% | 0% |
| Total intracranial volume, mean ± SD | 1428 ± 145 | 1420 ± 148 | 1434 ± 146 |
| Total grey matter volume, mean ± SD | 644 ± 66 | 639 ± 70 | 647 ± 65 |
| Total white matter volume, mean ± SD | 508 ± 62 | 502 ± 61 | 513 ± 64 |

*The demographics for the normosmic controls are the same as presented in Table 1.

The patient groups, separated into quantitative OD and parosmia and grouped together, scored significantly lower on the Sniffin’ Sticks test than the controls (all comparisons *p* < 0.001). Patients with parosmia scored slightly higher on olfactory function than patients with quantitative OD (*p*=0.05). There were no significant differences in total intracranial volume or total white matter volume between groups. Patients, both grouped and separated into their respective OD groups, had lower total grey matter volume compared with controls (*p*=0.006 for grouped OD, *p*=0.020 for patients with quantitative OD, and *p*=0.025 for patients with parosmia). All of these comparisons remained significant after adjusting for age and sex: *p* < 0.001, *p*=0.001, and *p*=0.001, respectively.

Appendix B: Supplementary materials GMD

Tables S2 – S6 contain the results from the GMD analysis with the two patients with no visible OBs removed. Overall, the results were similar to those from the full patient sample, with a few exceptions that are discussed here.

Whereas in the full patient sample no clusters passed the FWE cluster-forming threshold (*p=*0.05) in the whole-brain analysis, two small clusters were found in the comparison between patients with quantitative OD and controls: one in the right medial superior frontal gyrus and one in the left inferior parietal gyrus (Table S2).

**Table S2.** Brain regions with lower grey matter density for grouped COVID-19 OD patients (n=46) compared to normosmic controls (n=49) in whole-brain analysis – excluding patients with no visible OBs*

| Brain region | Cluster size  (voxels) | Hemisphere | Peak voxel MNI coordinates | | | Z-score |
| --- | --- | --- | --- | --- | --- | --- |
|  |  |  | X | Y | Z |  |
| Superior frontal gyrus, medial | 22 | R | 2 | 56 | 12 | 5.38 |
| Inferior parietal gyrus | 33 | L | -54 | -30 | 45 | 5.24 |

*Significant at FWE corrected threshold (*p* < 0.05), cluster extent threshold k=20.

Regarding the ROI analysis (still using our exploratory threshold), within the comparison between patients with quantitative OD and normosmic controls (Table S4), the clusters in the left medial orbital gyrus and dorsolateral superior frontal gyrus became part of a large cluster extending into the medial orbital superior frontal gyrus and gyrus rectus (k=603, Z=4.26). Once more, in the comparison between patients with quantitative OD and normosmic controls, a cluster in the right putamen appeared that was not present in the comparison with the full patient sample (k=195, Z=3.48). Finally, whereas the comparison between the two patient groups with the full patient sample showed no differences, the current analysis showed lower GMD in patients with quantitative OD compared to those with parosmia in the left nucleus accumbens (k=47, Z=3.71) and the right anterior insula (k=65, Z=3.66) (Table S6).

**Table S3.** Brain regions with lower grey matter density for grouped COVID-19 OD patients (n=46) compared to normosmic controls (n=49) within ROIs (exploratory analysis*) – excluding patients with no visible OBs.

| Brain region | Cluster size  (voxels) | Hemisphere | Peak voxel MNI coordinates | | | Z-score |
| --- | --- | --- | --- | --- | --- | --- |
|  |  |  | X | Y | Z |  |
| Anterior orbital gyrus^1^ | 69 | R | 17 | 59 | -14 | 4.59 |
| Middle frontal gyrus^2^ | 82 | R | 45 | 53 | -12 | 4.02 |
| Anterior orbital gyrus^3^ | 418 | L | -20 | 59 | -14 | 3.98 |
| Superior frontal gyrus, medial orbital |  | L | -3 | 63 | -15 | 3.81 |
| Superior frontal gyrus, medial orbital |  | L | -8 | 54 | -21 | 3.77 |
| Nucleus accumbens^4^ | 68 | R | 11 | 12 | -12 | 3.74 |
| Caudate nucleus | 61 | R | 6 | 12 | 0 | 3.72 |
| Olfactory cortex^5^ | 54 | L | -15 | 12 | -14 | 3.58 |

*Results presented are clusters with an extent threshold equal to the expected number of voxels per cluster; k=43 at a cluster-forming threshold of *p*=0.001 (uncorrected).

^1^Cluster extends into the right dorsolateral superior frontal gyrus.

^2^Cluster extends into the right anterior orbital gyrus and IFG pars orbitalis.

^3^Cluster extends into the left dorsolateral superior frontal gyrus, gyrus rectus, medial orbital gyrus and middle frontal gyrus.

^4^Cluster extends into the right gyrus rectus and olfactory cortex.

^5^Cluster extends into the left gyrus rectus, nucleus accumbens and putamen.

**Table S4.** Brain regions with lower grey matter density for COVID-19 OD patients with quantitative OD (n=20) compared to normosmic controls (n=49) within ROIs (exploratory analysis*) – excluding patients with no visible OBs.

| Brain region | Cluster size  (voxels) | Hemisphere | Peak voxel MNI coordinates | | | Z-score |
| --- | --- | --- | --- | --- | --- | --- |
|  |  |  | X | Y | Z |  |
| Anterior cingulate cortex, pregenual**^1^ | 104 | R | 2 | 54 | 11 | 5.28 |
| Superior frontal gyrus, medial |  | L | 3 | 50 | 18 | 3.83 |
| Superior frontal gyrus, medial orbital***^,2^ | 603 | L | -3 | 63 | -14 | 4.26 |
| Medial orbital gyrus |  | L | -9 | 56 | -21 | 4.04 |
| Superior frontal gyrus, dorsolateral |  | L | -23 | 66 | -6 | 3.81 |
| Anterior orbital gyrus^3^ | 64 | R | 17 | 60 | -14 | 3.98 |
| Nucleus accumbens**^,4^ | 545 | R | 12 | 11 | -12 | 3.97 |
| Nucleus accumbens |  | L | -5 | 8 | -6 | 3.86 |
| Olfactory cortex |  | L | -15 | 11 | -14 | 3.67 |
| Anterior orbital gyrus^5^ | 78 | R | 44 | 53 | -14 | 3.94 |
| Medial orbital gyrus | 78 | R | 14 | 39 | -20 | 3.94 |
| Gyrus rectus |  | R | 8 | 33 | -27 | 3.55 |
| Putamen | 195 | R | 26 | 8 | -6 | 3.48 |

*Results presented are clusters with an extent threshold equal to the expected number of voxels per cluster; k=42 at a cluster-forming threshold of *p*=0.001 (uncorrected).

**Peak voxel significant at *p*_fwe_ < 0.05.

***Significant at cluster-level *p*_fwe_ < 0.05.

^1^Cluster extends into the left pregenual anterior cingulate cortex.

^2^Cluster extends into the left anterior orbital gyrus and gyrus rectus.

^3^Cluster extends into the dorsolateral and medial orbital superior frontal gyrus.

^4^Cluster extends into the left gyrus rectus and putamen and the right caudate nucleus and olfactory cortex.

^5^Cluster extends into the right middle frontal gyrus.

**Table S5.** Brain regions with lower grey matter density for COVID-19 OD patients with parosmia (n=26) compared to normosmic controls (n=49) within ROIs (exploratory analysis*) – excluding patient with no visible OBs.

| Brain region | Cluster size  (voxels) | Hemisphere | Peak voxel MNI coordinates | | | Z-score |
| --- | --- | --- | --- | --- | --- | --- |
|  |  |  | X | Y | Z |  |
| Anterior orbital gyrus^1,2^ | 57 | L | -21 | 59 | -14 | 3.55 |

*Results presented are clusters with an extent threshold equal to the expected number of voxels per cluster; k=40 at a cluster-forming threshold of *p*=0.001 (uncorrected).

^1^Cluster extends into the left middle frontal gyrus.

^2^When controlling for Sniffin’ Sticks test score, this cluster no longer appears.

**Table S6.** Brain regions with lower grey matter density for COVID-19 OD patients with quantitative OD (n=20) compared to COVID-19 OD patients with parosmia (n=26) within ROIs (exploratory analysis*) – excluding patient with no visible OBs.

| Brain region | Cluster size  (voxels) | Hemisphere | Peak voxel MNI coordinates | | | Z-score |
| --- | --- | --- | --- | --- | --- | --- |
|  |  |  | X | Y | Z |  |
| Nucleus accumbens (Nacc)^1^ | 47 | L | -3 | 6 | -6 | 3.71 |
| Insula (anterior) | 65 | R | 41 | 5 | 6 | 3.66 |

**Results presented are clusters with an extent threshold equal to the expected number of voxels per cluster; k=37 at a cluster-forming threshold of *p*=0.001 (uncorrected).

^1^Cluster extends into the left olfactory cortex.

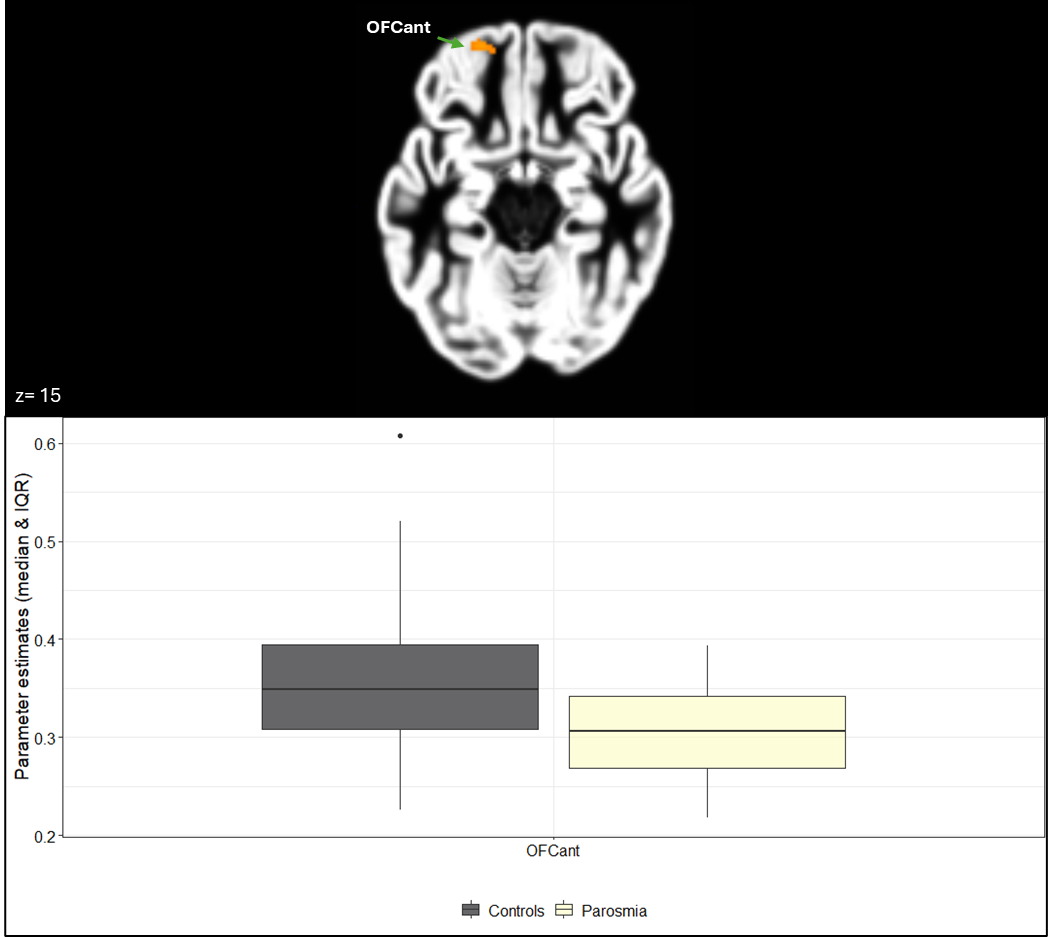

**Fig. S1.** Colour-coded T-maps thresholded at T=3.21 (cluster-forming threshold *p*=0.001 (uncorrected), cluster extent threshold k=41 (expected number of voxels)) showing lower GMD in COVID-19 patients with quantitative OD versus normosmic controls within predefined ROIs, overlaid onto the mean sample GM image (top) and boxplot of associated average parameter estimates (bottom). OFCant = anterior orbital gyrus. The lower and upper whiskers of the boxplots represent the smallest and largest values within 1.5 times the interquartile range (IQR), respectively. Data points beyond 1.5 times the interquartile range are plotted individually.

Appendix C: Supplementary materials odour-induced brain activation

**Table S7.** Participant demographics – fMRI analysis

|  | Controls  (n=46) | Grouped OD  (n=46) |  |  |
| --- | --- | --- | --- | --- |
| Patient subgroup |  |  | Quantitative OD  (n=20) | Qualitative OD  (n=26) |
| Age, mean ± SD (range) | 40.3 ± 13.2  (22-60) | 47.2 ± 9.6  (19-60) | 44.1 ± 11.1  (19-58) | 49.6 ± 7.6  (31-60) |
| Gender, female/male | 22/24 | 37/9 | 15/5 | 22/4 |
| Sniffin’ Sticks score, mean ± SD (range) | 34.5 ± 2.6  (31-41.5) | 22.8 ± 6.9  (6-33.25) | 20.1 ± 8.4  (6-29.5) | 24.9 ± 4.5  (17.5-33.25) |
| Days since SARS-CoV-2 diagnosis, mean ± SD (range) | N/A | 445 ± 463  (86-1500) | 573 ± 557  (90-1500) | 347 ± 356  (86-1212) |
| Severity of infection |  |  |  |  |
| *Non-symptomatic/mild* | N/A | 21.7% | 25.0% | 19.2% |
| *Moderate* | N/A | 65.2% | 50.0% | 76.9% |
| *Severe (hospital admission)* | N/A | 13.0% | 25.0% | 3.8% |
| *Critical (intensive care admission)* | N/A | 0% | 0% | 0% |
| Total intracranial volume, mean ± SD | 1480 ± 159 | 1434 ± 148 | 1432 ± 157 | 1436 ± 145 |
| Total grey matter volume, mean ± SD | 688 ± 83 | 645 ± 68 | 644 ± 73 | 646 ± 65 |
| Total white matter volume, mean ± SD | 522 ± 73 | 509 ± 62 | 503 ± 62 | 514 ± 63 |

The patient groups, separated into quantitative OD and parosmia and grouped together, scored significantly lower on the Sniffin’ Sticks test than the controls (all comparisons *p* < 0.001). Patients with parosmia scored slightly higher on olfactory function than patients with quantitative OD (*p*=0.03). There were no significant differences in total intracranial volume or total white matter volume between groups. Patients, both grouped and separated into their respective OD groups, had lower total grey matter volume compared with controls (*p*=0.008 for grouped OD, *p*=0.037 for patients with quantitative OD, and *p*=0.022 for patients with parosmia). All of these comparisons remained significant after adjusting for age and sex: *p* < 0.001, *p*=0.002, and *p*=0.001, respectively.

**Table S8.** Odour-induced brain activation of normosmic controls (n=46) within ROIs.*

| Contrast | Brain region | Cluster size  (voxels) | Hemisphere | Peak voxel MNI coordinates | | | Z-score |
| --- | --- | --- | --- | --- | --- | --- | --- |
|  |  |  |  | X | Y | Z |  |
| OdourTot > rest | Insula (anterior)^1^ | 2489 | R | 35 | 23 | -4 | Inf |
|  | Insula (anterior) |  | R | 41 | 18 | 3 | 7.47 |
|  | Insula (anterior) |  | R | 48 | 16 | -2 | 7.2 |
|  | Insula (anterior)^2^ | 2523 | L | -31 | 25 | -2 | 7.66 |
|  | Hippocampus |  | L | -22 | -25 | -8 | 7.62 |
|  | Temporal pole: superior  temporal gyrus |  | L | -57 | 12 | -2 | 7.38 |
|  | Lateral geniculate^3^ | 215 | R | 22 | -27 | -6 | 6.75 |
|  | Lingual gyrus |  | R | 15 | -36 | -6 | 5.55 |
|  | Hippocampus |  | R | 24 | -36 | 0 | 4.4 |
|  | Anterior cingulate cortex, supracallosal^4^ | 959 | L | -4 | 27 | 27 | 6.73 |
|  | Anterior cingulate cortex, supracallosal |  | R | 11 | 33 | 22 | 6.11 |
|  | Anterior cingulate cortex, supracallosal |  | R | 6 | 14 | 25 | 5.9 |
| OdourTot > blank | Hippocampus | 198 | L | -22 | -21 | -13 | 4.98 |
|  | Amygdala |  | L | -26 | -1 | -19 | 4.9 |
|  | Parahippocampal gyrus |  | L | -13 | -6 | -17 | 4.86 |

*Cluster-forming threshold of *p*=0.001 (uncorrected). Presented results are significant at cluster-level *p*_fwe_ < 0.05.

^1^Cluster extends into the right amygdala, caudate nucleus, gyrus rectus, hippocampus, IFG pars orbitalis, medial and posterior orbital gyri, nucleus accumbens, olfactory cortex, parahippocampal gyrus, putamen and temporal pole: superior temporal gyrus.

^2^Cluster extends into the left amygdala, caudate nucleus, dorsolateral superior frontal gyrus, gyrus rectus, IFG pars orbitalis, lateral geniculate, medial and posterior orbital gyri, middle frontal gyrus, nucleus accumbens, olfactory cortex, parahippocampal gyrus and putamen.

^3^Cluster extends into the right parahippocampal gyrus.

^4^Cluster extends into the pregenual anterior cingulate cortex (bilaterally).

**Table S9.** Odour-induced brain activation of grouped COVID-19 OD patients (n=46) within ROIs.*

| Contrast | Brain region | Cluster size  (voxels) | Hemisphere | Peak voxel MNI coordinates | | | Z-score |
| --- | --- | --- | --- | --- | --- | --- | --- |
|  |  |  |  | X | Y | Z |  |
| OdourTot > rest | Insula (anterior)^1^ | 2759 | L | -31 | 23 | 0 | Inf |
|  | Insula (anterior) |  | L | -39 | 16 | -2 | 7.27 |
|  | Temporal pole: superior  temporal gyrus |  | L | -48 | 12 | -6 | 7.01 |
|  | Insula (anterior)^2^ | 2706 | R | 39 | 20 | 3 | 7.63 |
|  | Insula (anterior) |  | R | 35 | 25 | -4 | 7.61 |
|  | Insula (anterior) |  | R | 48 | 14 | -6 | 7.42 |
|  | Hippocampus | 232 | L | -22 | -27 | -6 | 7.61 |
|  | Parahippocampal gyrus |  | L | -15 | -40 | -4 | 5.57 |
|  | Fusiform gyrus |  | L | -33 | -45 | -11 | 3.36 |
|  | Hippocampus^3^ | 276 | R | 22 | -25 | -11 | 6.77 |
|  | Hippocampus |  | R | 19 | -30 | -4 | 6.42 |
|  | Lingual gyrus |  | R | 22 | -45 | -6 | 6.30 |
|  | Anterior cingulate cortex, supracallosal^4^ | 1118 | L | -11 | 23 | 29 | 6.74 |
|  | Anterior cingulate cortex, supracallosal |  | R | 11 | 29 | 27 | 6.56 |
|  | Anterior cingulate cortex, supracallosal |  | R | 9 | 18 | 27 | 6.10 |
| OdourTot > blanks | Amygdala^5^ | 101 | R | 22 | -3 | -13 | 5.91 |
|  | Amygdala^6^ | 121 | L | -20 | -3 | -15 | 4.93 |
|  | Amygdala |  | L | -28 | -1 | -17 | 4.68 |

*Cluster-forming threshold of *p*=0.001 (uncorrected). Presented results are significant at cluster-level *p*_fwe_ < 0.05.

^1^Cluster extends into the left amygdala, caudate nucleus, dorsolateral superior frontal gyrus, gyrus rectus, hippocampus, IFG pars orbitalis, medial and posterior orbital gyri, middle frontal gyrus, nucleus accumbens, olfactory cortex and putamen.

^2^Cluster extends into the right amygdala, caudate nucleus, gyrus rectus, hippocampus, IFG pars orbitalis, medial and posterior orbital gyri, nucleus accumbens, olfactory cortex, putamen and temporal pole: superior and middle temporal gyri.

^3^Cluster extends into the right parahippocampal gyrus.

^4^Cluster extends into the pregenual anterior cingulate cortex (bilaterally).

^5^Cluster extends into the right hippocampus and parahippocampal gyrus.

^6^Cluster extends into the left hippocampus.

**Table S10.** Odour-induced brain activation of COVID-19 OD patients with quantitative OD (n=20) within ROIs.*

| Contrast | Brain region | Cluster size  (voxels) | Hemisphere | Peak voxel MNI coordinates | | | Z-score |
| --- | --- | --- | --- | --- | --- | --- | --- |
|  |  |  |  | X | Y | Z |  |
| OdourTot > rest | IFG pars orbitalis^1^ | 1661 | L | -46 | 20 | -4 | 5.38 |
|  | Insula (anterior) |  | L | -31 | 25 | 0 | 5.33 |
|  | Insula (anterior) |  | L | -37 | 16 | 7 | 5.23 |
|  | Insula (anterior)^2^ | 1604 | R | 45 | 23 | -6 | 5.21 |
|  | Insula (anterior) |  | R | 37 | 20 | 5 | 5.16 |
|  | Olfactory cortex |  | R | 15 | 12 | -19 | 5.16 |
|  | Hippocampus^3^ | 142 | R | 24 | -25 | -11 | 5.11 |
|  | Hippocampus |  | R | 17 | -34 | 3 | 4.38 |
|  | Hippocampus |  | R | 19 | -32 | -6 | 4.37 |
|  | Anterior cingulate cortex, supracallosal^4^ | 741 | L | -4 | 10 | 29 | 5.08 |
|  | Anterior cingulate cortex, supracallosal |  | L | -11 | 23 | 29 | 4.67 |
|  | Anterior cingulate cortex, supracallosal |  | L | -2 | 20 | 22 | 4.44 |
|  | Hippocampus^5^ | 162 | L | -22 | -25 | -8 | 4.7 |
|  | Parahippocampal gyrus |  | L | -15 | -40 | -6 | 4.21 |
|  | Hippocampus |  | L | -17 | -36 | 0 | 4.19 |
| OdourTot > blanks | N/A | N/A | N/A | N/A | N/A | N/A | N/A |

*Cluster-forming threshold of *p*=0.001 (uncorrected). Presented results are significant at cluster-level *p*_fwe_ < 0.05.

^1^Cluster extends into the left putamen, caudate nucleus, olfactory cortex, amygdala, parahippocampal gyrus, nucleus accumbens, gyrus rectus, IFG pars orbitalis, middle frontal gyrus, temporal pole: superior temporal gyrus and medial and posterior orbital gyri.

^2^Cluster extends into the right putamen, caudate nucleus, olfactory cortex, amygdala, gyrus rectus, hippocampus, nucleus accumbens, IFG pars orbitalis and medial and posterior orbital gyri.

^3^Cluster extends into the right parahippocampal gyrus and lateral geniculate.

^4^Cluster extends into the right supracallosal and bilateral pregenual anterior cingulate cortex.

^5^Cluster extends into the left lateral geniculate.

**Table S11.** Odour-induced brain activation of COVID-19 OD patients with parosmia (n=26) within ROIs.*

| Contrast | Brain region | Cluster size  (voxels) | Hemisphere | Peak voxel MNI coordinates | | | Z-score |
| --- | --- | --- | --- | --- | --- | --- | --- |
|  |  |  |  | X | Y | Z |  |
| OdourTot > rest | Insula (anterior)^1^ | 1198 | R | 39 | 18 | 0 | 6.33 |
|  | Insula (anterior) |  | R | 35 | 25 | -2 | 5.86 |
|  | IFG pars orbitalis |  | R | 52 | 18 | -6 | 5.69 |
|  | Lateral geniculate^2^ | 125 | L | -22 | -27 | -6 | 6.11 |
|  | Parahippocampal gyrus |  | L | -15 | -40 | -4 | 3.84 |
|  | IFG pars orbitalis^3^ | 1714 | L | -39 | 20 | -6 | 6.02 |
|  | Temporal pole: superior  temporal gyrus |  | L | -50 | 12 | -6 | 5.92 |
|  | Insula (anterior) |  | L | -33 | 23 | 0 | 5.88 |
|  | Caudate nucleus^4^ | 457 | R | 15 | 18 | 0 | 5.7 |
|  | Caudate nucleus |  | R | 13 | 12 | 5 | 5.32 |
|  | Putamen |  | R | 19 | 5 | 7 | 4.54 |
|  | Lingual gyrus^5^ | 166 | R | 19 | -45 | -6 | 5.21 |
|  | Lateral geniculate |  | R | 22 | -23 | -8 | 5.19 |
|  | Hippocampus |  | R | 17 | -36 | 0 | 3.93 |
|  | Anterior cingulate cortex, supracallosal^6^ | 535 | R | 11 | 31 | 25 | 4.84 |
|  | Anterior cingulate cortex, supracallosal |  | R | 9 | 18 | 27 | 4.82 |
|  | Anterior cingulate cortex, supracallosal |  | L | 0 | 27 | 20 | 4.81 |
| OdourTot > blanks | Hippocampus^7^ | 61 | R | 19 | -3 | -15 | 4.76 |
|  | Amygdala^8^ | 80 | L | -24 | 1 | -17 | 4.47 |
|  | Amygdala |  | L | -15 | -6 | -17 | 4.2 |

*Cluster-forming threshold of *p*=0.001 (uncorrected). Presented results are significant at cluster-level *p*_fwe_ < 0.05.

^1^Cluster also extends into the right gyrus rectus, medial and posterior orbital gyri, olfactory cortex, putamen, and temporal pole: superior temporal gyrus.

^2^Cluster extends into the left hippocampus.

^3^Cluster extends into the left amygdala, caudate nucleus, dorsolateral superior frontal gyrus, gyrus rectus, medial and posterior orbital gyri, middle frontal gyrus, nucleus accumbens, putamen and Rolandic uperculum,

^4^Cluster extends into the right nucleus accumbens.

^5^Cluster extends into the right parahippocampal gyrus.

^6^Cluster extends into the pregenual anterior cingulate cortex (bilaterally).

^7^Cluster extends into the right amygdala.

^8^Cluster extends into the left hippocampus.

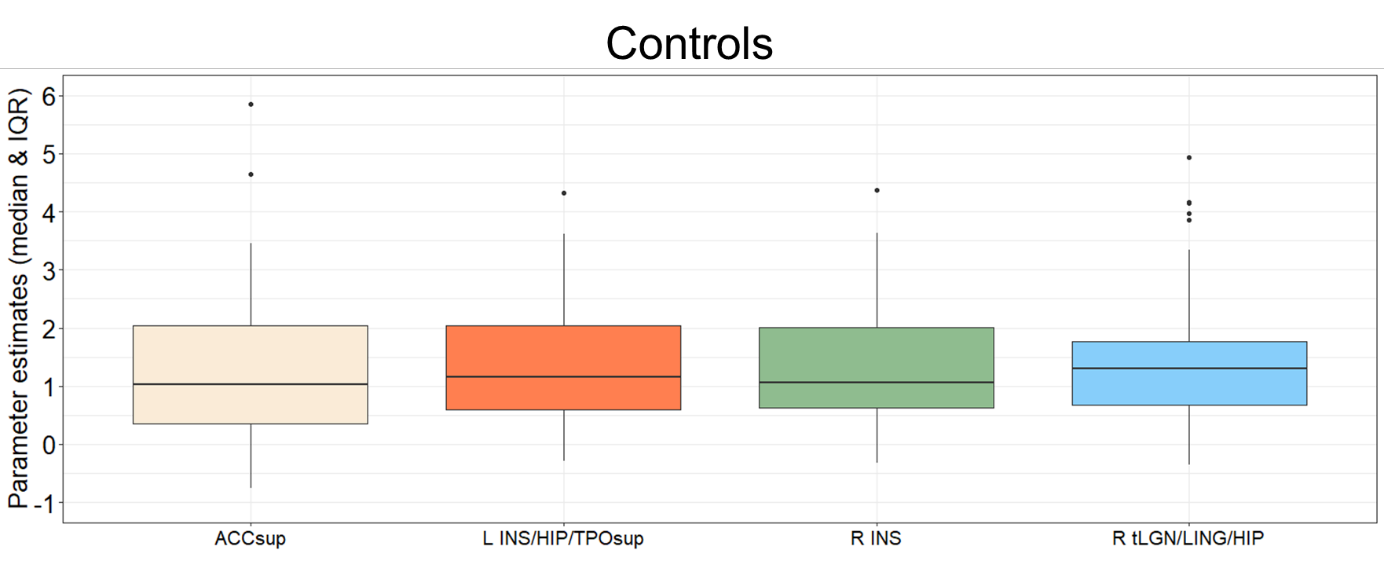

**Fig. S2.** Boxplot of average parameter estimates of odour-induced brain activation (OdourTot > rest contrast) in normosmic controls (n=46). ACCsup = anterior cingulate cortex, supracallosal; INS = insula (anterior); HIP = hippocampus; TPOsup = Temporal pole: superior temporal gyrus; tLGN= lateral geniculate; LING = lingual gyrus. The lower and upper whiskers of the boxplots represent the smallest and largest values within 1.5 times the interquartile range (IQR), respectively. Data points beyond 1.5 times the interquartile range are plotted individually.

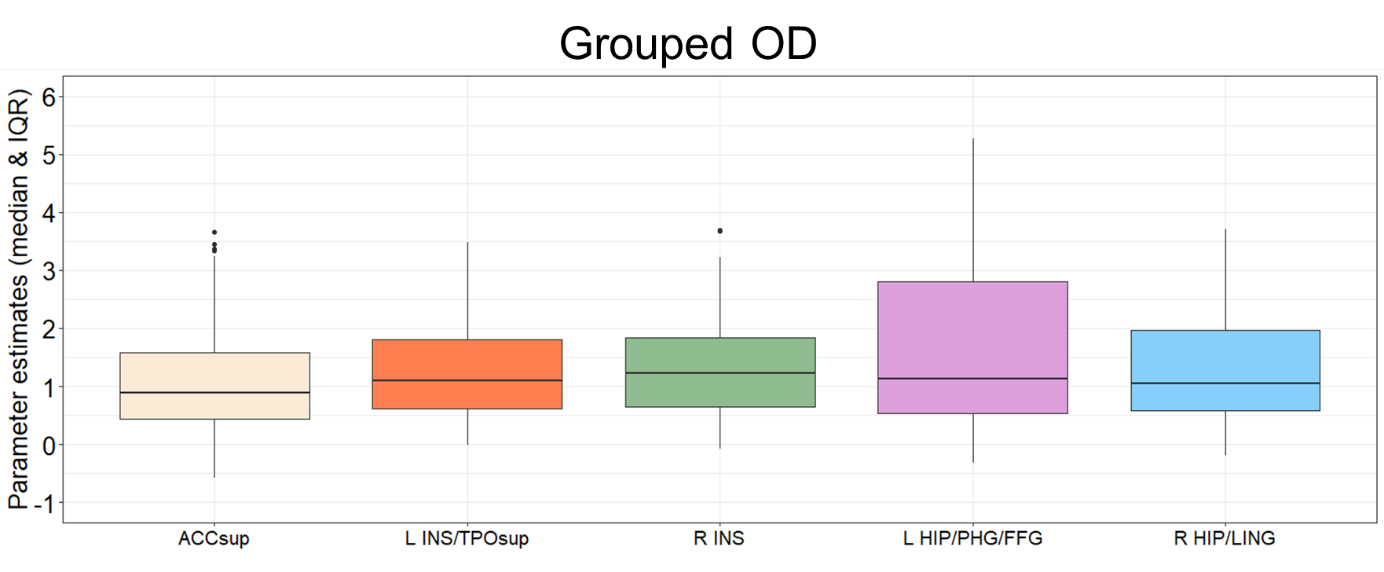

**Fig. S3.** Boxplot of average parameter estimates of odour-induced brain activation (OdourTot > rest contrast) in grouped COVID-19 OD patients (n=46). ACCsup = anterior cingulate cortex, supracallosal; INS = insula (anterior); TPOsup = Temporal pole: superior temporal gyrus; HIP = hippocampus; PHG = parahippocampal gyrus; FFG = fusiform gyrus; LING = lingual gyrus. The lower and upper whiskers of the boxplots represent the smallest and largest values within 1.5 times the interquartile range (IQR), respectively. Data points beyond 1.5 times the interquartile range are plotted individually.

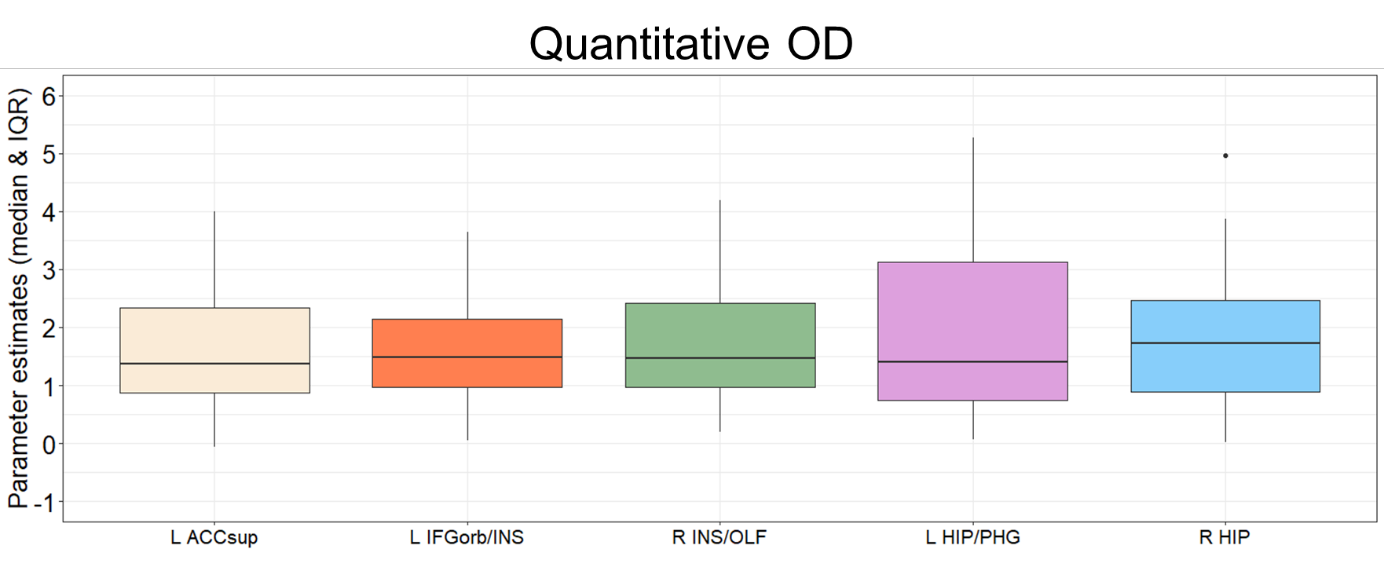

**Fig. S4.** Boxplot of average parameter estimates of odour-induced brain activation (OdourTot > rest contrast) in COVID-19 OD patients with quantitative OD (n=20). ACCsup = anterior cingulate cortex, supracallosal; IFGorb = IFG pars orbitalis; INS = insula (anterior); OLF = olfactory cortex; HIP = hippocampus; PHG = parahippocampal gyrus. The lower and upper whiskers of the boxplots represent the smallest and largest values within 1.5 times the interquartile range (IQR), respectively. Data points beyond 1.5 times the interquartile range are plotted individually.

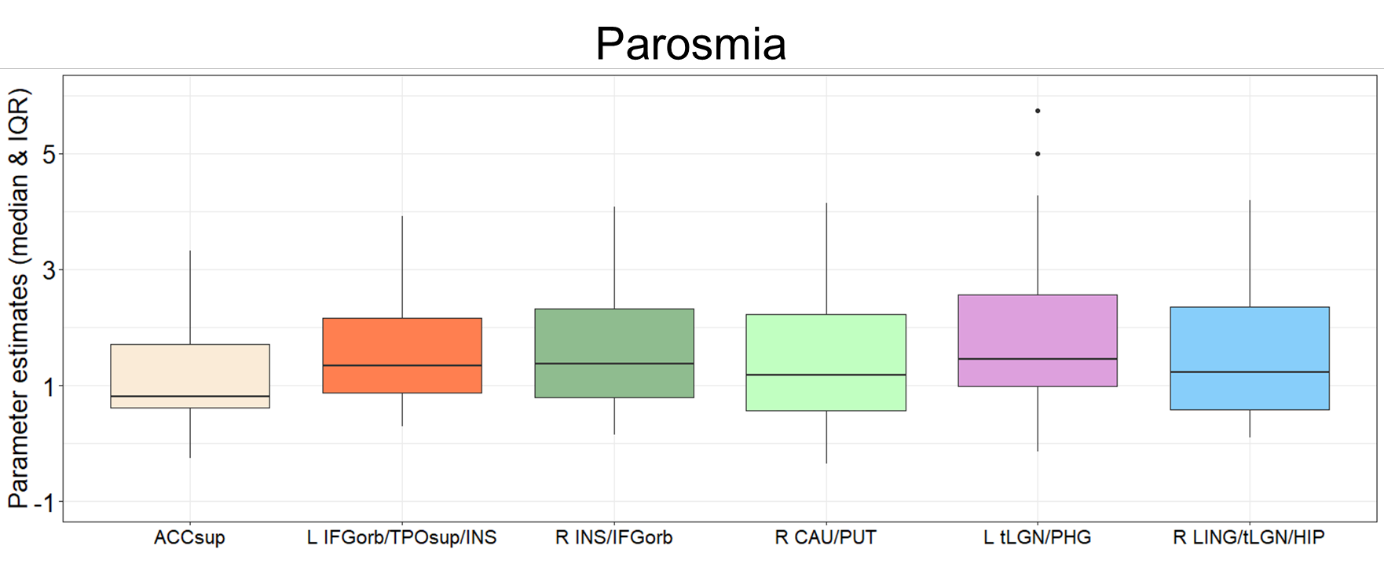

**Fig. S5.** Boxplot of average parameter estimates of odour-induced brain activation (OdourTot > rest contrast) in COVID-19 patients with parosmia (n=26). ACCsup = anterior cingulate cortex, supracallosal; IFGorb = IFG pars orbitalis; TPOsup = Temporal pole: superior temporal gyrus; INS = insula (anterior); CAU = caudate; PUT = putamen; tLGN= lateral geniculate; PHG = parahippocampal gyrus; LING = lingual gyrus; HIP = hippocampus. The lower and upper whiskers of the boxplots represent the smallest and largest values within 1.5 times the interquartile range (IQR), respectively. Data points beyond 1.5 times the interquartile range are plotted individually.

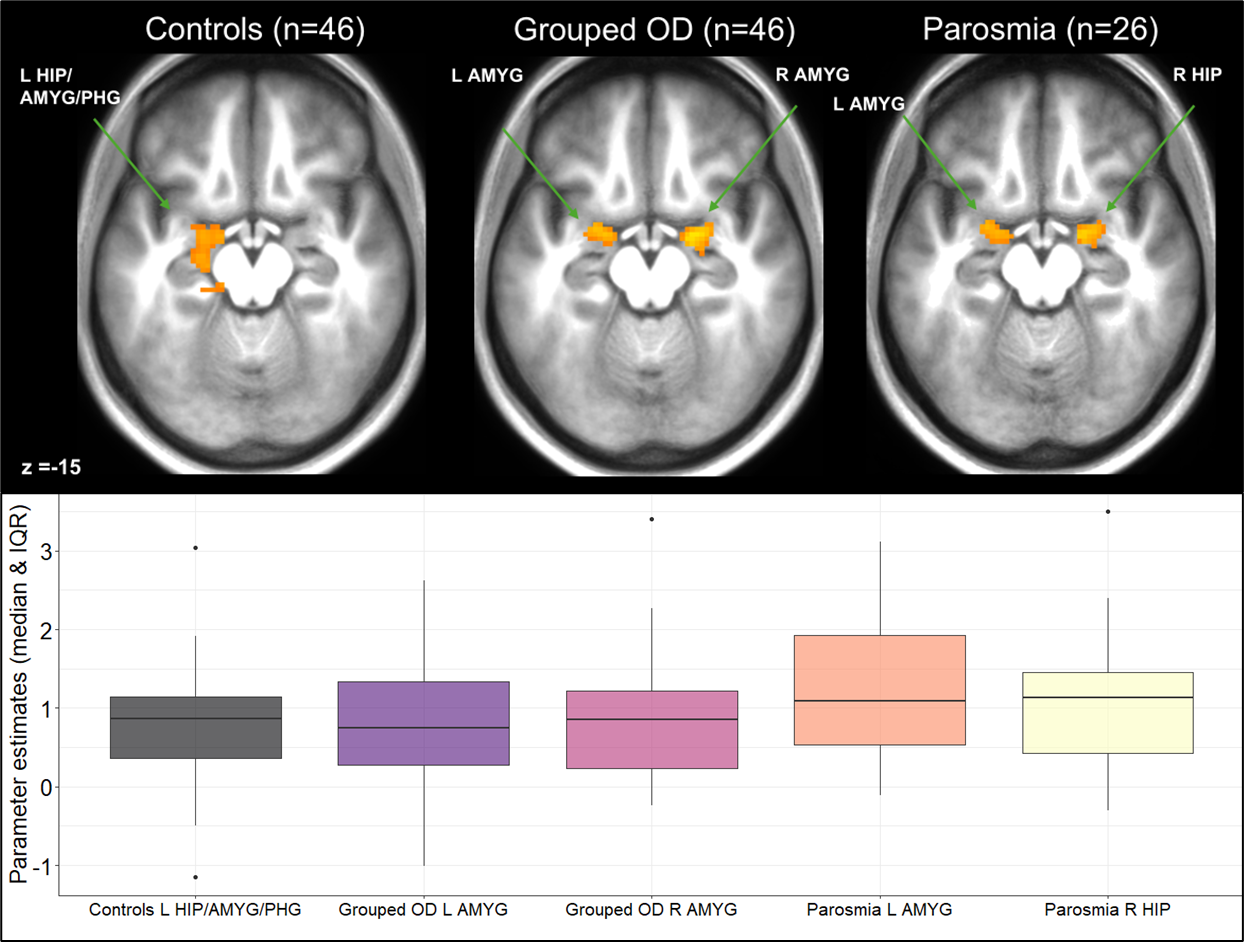

**Fig. S6.** Color-coded T-maps thresholded at T=3.29 (cluster-forming threshold *p*=0.001 (uncorrected)), showing odour-induced brain activation (OdourTot > blanks contrast) per group, overlaid onto mean respective anatomical image (top). Shown activations are significant at cluster-level *p*_fwe_ < 0.05. Boxplot of associated parameter estimates (bottom). HIP = hippocampus; AMYG = amygdala; PHG = parahippocampal gyrus. The lower and upper whiskers of the boxplots represent the smallest and largest values within 1.5 times the interquartile range (IQR), respectively. Data points beyond 1.5 times the interquartile range are plotted individually.
